# Spatially resolved T cell receptor tracking reveals γδT cell localization to tumor-rich regions in high-risk neuroblastoma: A Report from the Children’s Oncology Group

**DOI:** 10.64898/2026.06.10.26354144

**Authors:** Yiyue Jiang, Wenbao Yu, Yanan Wang, Anusha Thadi, Stephanie Pedersen, Jenna Eagles, Arlene Naranjo, Natalie Collins, Steven G. DuBois, Rochelle Bagatell, Brian D. Crompton, Kai Tan, Trevor J. Pugh

## Abstract

High-risk neuroblastoma (HRNB) is a leading cause of pediatric cancer death. Current therapies center on intensive multimodal treatment including anti-GD2 therapy, with growing interest in harnessing T cell-mediated immunity. How T cells and their receptors (T-cell receptors, TCRs) are spatially organized and function within tumors remains poorly defined. To assess whether intratumoral location influences clonotype-specific T cell states, we profiled TCR repertoires across blood and tumor samples from 37 patients with HRNB using longitudinal bulk TCR sequencing. In a nested subset of 5 patients with paired pre- and post-therapy tumors, we integrated spatial transcriptomics with *in situ* TCR profiling. Across all tumors, T and B cells preferentially co-localized in immune-rich regions and showed reduced proximity to neuroblast cells. Despite this compartmentalized architecture, γδT cells were more evenly distributed across tumor sections and showed greater proximity to neuroblast-rich regions than other T cell subsets. Within TCR clonotypes, spatial location was associated with distinct transcriptional states, with immune-rich regions supporting more progenitor-like programs. These findings identify spatial context as a key determinant of phenotype clonotype-specific T cell phenotype and highlight γδT cells cells as a spatially distinct population with potential roles in neuroblastoma tumor-immune interactions.

## Introduction

Neuroblastoma is the most common extracranial solid tumor in children^1^, arising from neural crest-derived sympathoadrenal progenitors and most frequently originating in the adrenal medulla^2^. Approximately half of newly diagnosed patients have high-risk neuroblastoma (HRNB), most commonly defined by metastatic disease diagnosed after 18 months of age, or by amplification of the oncogene *MYCN* in locoregional tumors^3^. Standard-of-care (SOC) therapy occurs over 18 months and includes induction chemotherapy, surgical resection of the primary tumor, consolidation with high dose chemotherapy, autologous stem-cell transplant and external beam radiotherapy, followed by post-consolidation therapy with anti-GD2 monoclonal antibody therapy^4^. Together, these intensive therapies not only reduce tumor burden but also profoundly reshape the immune system through lymphodepletion, antigen release, and subsequent immune reconstitution^5,6^. Despite these interventions, 5-year event-free survival remains just over 50%^7^, underscoring the need to better understand the biological and immune factors that limit durable disease control in HRNB.

T cells are central mediators of adaptive antitumor immunity^8^. Through V(D)J recombination, they generate diverse T cell receptors (TCRs) defined by unique complementarity-determining region 3 (CDR3) sequences that confer antigen specificity^9^. In neuroblastoma, cytotoxic αβ CD8+ T cells are frequently detected but functionally constrained as low tumor mutational burden limits neoantigen availability, and downregulation of MHC class I expression impairs antigen presentation^10^. These features have drawn attention to γδT cells, which recognize targets in an MHC-independent manner and have been identified in HRNB tumors^11^. Nevertheless, the overall T cell infiltrate remains variable and functionally limited, consistent with an immunologically cold tumor microenvironment^12^.

Frontline clinical trial samples provide an opportunity to examine how therapeutic interventions shape T cell immunity over time. ANBL1531 is a Phase 3 randomized trial (NCT03126916) evaluating ^131^I-metaiodobenzylguanidine (^131^I-MIBG) administered before induction cycle 4 in combination with standard therapy compared with standard therapy alone. ^131^I-MIBG is a norepinephrine analog that gets selectively taken up by neuroblastoma cells^13^. It delivers targeted radiation to sites of disease and has demonstrated activity in relapsed HRNB^13^.

Recent single-cell and spatial profiling studies have begun to define the architectural features of the neuroblastoma tumor microenvironment. Integrative analyses have revealed transcriptionally distinct tumor cell states organize along developmental gradients, with immune-excluded tumor-stroma interfaces enriched for macrophages and fibroblasts that physically limit T-cell infiltration, alongside therapy-associated expansion of pro-tumorigenic myeloid populations that further promote immune evasion^14–18^. In parallel, a spatial TCR profiling study in adult head and neck squamous cell carcinoma found that T cells sharing identical CDR3 sequences can occupy different tumor regions and exhibit divergent activation or exhaustion states^19^. While this study highlights the importance of local microenvironmental cues in shaping T-cell behavior, this raises the question of whether similar spatial regulation of T-cell fate operates in pediatric neuroblastoma.

Here, we sought to characterize how T-cell clonality, spatial organization, and phenotypic state evolve over time in HRNB. By integrating longitudinal TCR sequencing from tumor tissue, peripheral blood, and cell-free DNA (cfDNA) with spatially resolved *in situ* TCR and transcriptomic profiling, we examined how tumor-immune segregation and spatial context relate to T cell exhaustion and clonal behavior within the tumor microenvironment. Leveraging longitudinal samples collected across ANBL1531, our analysis captures tumor-immune dynamics across clinical timepoints during frontline therapy, without formal treatment-arm-specific comparisons.

## Results

### Longitudinal bulk TCR profiling defined repertoire dynamics

To profile the immune landscape of HRNB, we evaluated 118 blood and tumour samples from 37 patients enrolled on the ANBL1531 clinical trial, spanning up to 11 clinical timepoints per patient (Fig. 1a,b). This cohort included five patients with paired tumor samples, for whom treatment-arm assignment was released by ANBL1531. This enabled analysis of longitudinal TCR dynamics across patients and compartments as well as subsequent spatial transcriptomic and TCR mapping within tumor tissue.

**Figure 1.**
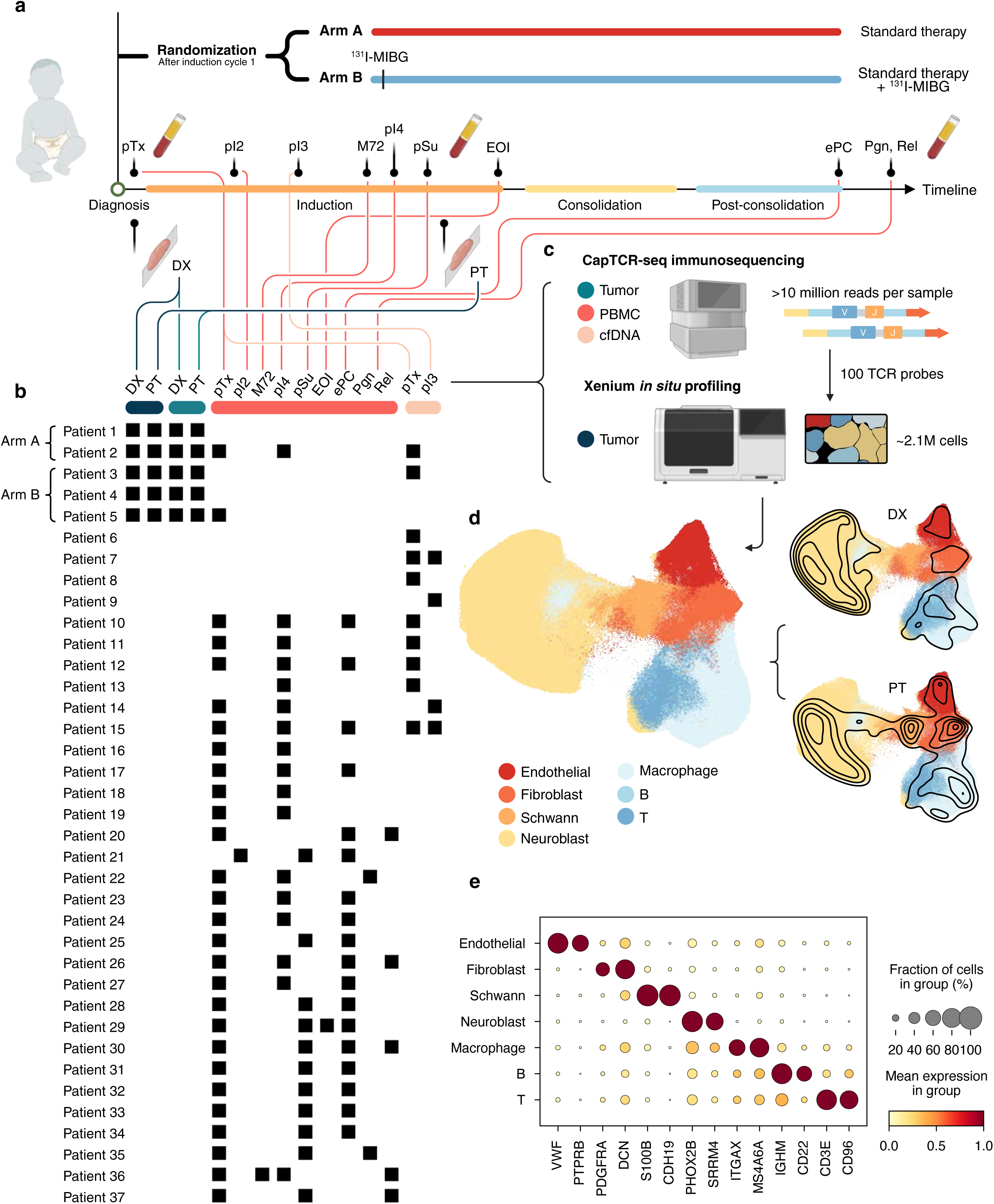
Cohort design and Xenium-based cellular atlas of primary HRNB tumors. **a**, Schematic of the ANBL1531 clinical trial timeline annotated with longitudinal sampling milestones. **b**, Overview of cohort structure, showing sample types, longitudinal timepoints for each patient group, and arm assignment for five patients. **c**, Schematic of experimental workflow applied to cohort specimens. **d**, UMAP of 2,072,978 integrated Xenium cells colored by annotated cell type (left), with Gaussian kernel density contour overlays for DX (upper right) and PT (lower right) samples shown separately. **e**, Dot plot of canonical markers for each of the seven major cell types. Dot size, fraction of cells expressing each gene; color, standardized mean expression. pTx, pre-treatment; DX, diagnostic biopsy; pI2-pI4, pre-induction cycles 2-4; M72, 72 h pre-131I-MIBG; pSu, pre-surgery; PT, surgical resection; EOI, end of induction; ePC, end of post-consolidation; Pgn, progression; Rel, relapse. Panels a-c were created with BioRender.com.

We performed bulk TCR CDR3 sequencing (CapTCR-seq)^20^ on formalin-fixed, paraffin-embedded (FFPE) tumor tissue (n = 10), peripheral blood mononuclear cells (PBMCs, n = 84), and cfDNA (n = 14) (median paired reads per sample: 27.5 million for PBMC, 35.6 million for tumor, 12.4 million for cfDNA, Fig. 1b,c). FFPE tumor samples consisted of paired diagnostic biopsies (DX) and post-chemotherapy surgical resections (PT) from five patients (n = 10 samples, 2 matched samples per patient). PBMC and cfDNA samples spanned longitudinal clinical milestones, including diagnosis, pre-induction cycles 2, 3 and 4, 72 hours post-^131^I-MIBG treatment, pre-surgical resection, end-of-induction, end-of-post-consolidation, progression/relapse (Fig. 1b). Across 108 bulk-sequenced samples, we identified 113,799 unique CDR3 clonotypes, with 90% detected in PBMCs, 9.5% in tumor tissue and 0.43% in cfDNA (Table 1).

**Table 1.** Summary of unique CDR3 clonotypes identified by bulk TCR sequencing.

| Sample type | Samples, n | $\gamma$ clonotypes | $\alpha/\delta$ clonotypes | $\beta$ clonotypes | Total clonotypes |
| --- | --- | --- | --- | --- | --- |
| Tumor | 10 | 2,000 | 6,207 | 2,564 | 10,771 |
| PBMC | 84 | 17,815 | 55,631 | 29,088 | 102,534 |
| cfDNA | 14 | 89 | 279 | 126 | 494 |
| All samples | 108 | 19,904 | 62,117 | 31,778 | 113,799 |

Clonotype counts are shown by sample type and TCR chain group across all 108 bulk-sequenced samples. $\alpha$ and $\delta$ chains are reported together because they were captured and analyzed as a combined $\alpha/\delta$ assay.

From these data, we assessed changes in TCR repertoire diversity across α, β and γ chains. Diversity of δ chain was not analyzed because the δ locus is embedded within the α locus^21^, precluding unambiguous assignment. cfDNA was also excluded due to low TCR clone count.

In PBMCs, repertoire diversity decreased after treatment initiation across α, β and γ chains. From pre-treatment to pre-induction cycle 4 to pre-surgery, mean diversity decreased from 980 to 245 to 120 for α, 491 to 226 to 151 for β, and 265 to 145 to 74 for γ. These decreases were significant across all chains (Holm-adjusted LMM contrasts: α, pre-treatment to pre-induction cycle 4 P<0.001, pre-treatment to pre-surgery P<0.001; β, P=0.010, P=0.004; γ, P=0.021, P=0.001) (Extended Data Fig. 1a-c). Diversity returned to baseline by end of post-consolidation (α, 980 to 780, P=0.405; β, 491 to 432, P=0.806; γ, 265 to 244, P=0.553), consistent with transient chemotherapy-induced lymphodepletion^22^.

In the 5 paired tumors sampled by biopsy at diagnosis and surgical resection, repertoire diversity decreased across all chains, although these changes were not statistically significant (diagnostic biopsy to surgical resection: α, 798 to 100, P=0.19; β, 311 to 80, P=0.20; γ, 221 to 65, P=0.15) (Extended Data Fig. 1d-f). Diversity changes also showed no consistent directionality by treatment arm (Arm A, SOC, Patients 1-2; Arm B, SOC + MIBG, Patients 3-5; two-sided Fisher’s exact test for direction of change: α, β and γ, P=1.0) (Extended Data Fig. 1 source data). Together, these data suggest that systemic and intratumoral TCR repertoires contract during therapy, with PBMC diversity recovering after treatment whereas tumor repertoire changes were more variable and not clearly associated with treatment arm.

To assess cross-compartment sharing of TCR clonotypes, we quantified overlap among available matched cfDNA, PBMC and tumor samples from 9 patients. This included PBMC-tumor and cfDNA-tumor comparisons in patients with available tumor material, and cfDNA-PBMC comparisons in patients with matched circulating samples.

In PBMC-tumor comparisons (n=2), 4.1%, 2.6%, and 12% of PBMC clonotypes were detected in tumors (ranges 2.6-5.6%, 1.3-3.9%, and 10-13%), compared to 4.4%, 5.0%, and 9.5% of tumor clonotypes detected in PBMCs (ranges 0.92-7.8%, 0.46-9.5%, and 4.8-14%).

In cfDNA-tumor comparisons (n=2), medians of 5.3%, 9.1%, and 30% of cfDNA clonotypes (TCRα/δ, TCRβ, and TCRγ, respectively) were detected in tumors (ranges 0-11%, 0-18%, and 26-33%), compared to 0.20%, 0.41%, and 0.97% of tumor clonotypes detected in cfDNA (ranges 0-0.41%, 0-0.83%, and 0.39-1.5%). Overall, cfDNA repertoires showed greater overlap with tumor-associated clonotypes than PBMC repertoires, particularly for TCRγ.

Consistent with the limited overlap between circulating compartments, cfDNA-PBMC comparisons (n=7) showed low overlap across chains. For TCRα/δ, 1.3% (range 0-11%) of cfDNA clonotypes were detected in PBMCs versus 0.02% (range 0-0.15%) in the reverse direction. For TCRβ, median overlap was 0 in both directions (ranges 0-9.1% and 0-0.17%). For TCRγ, overlap was asymmetric, with 11% (range 0-67%) of cfDNA clonotypes detected in PBMCs versus 0.28% (range 0-0.62%) in reverse.

### Spatial profiling identifies heterogeneous tumor-immune composition across timepoints

To spatially localize clonotypes identified by bulk TCR sequencing, we designed a Xenium panel containing 100 candidate TCR CDR3 sequences selected from longitudinal bulk repertoires (Extended Data Fig. 1g, Supplementary Table 1, Methods). We prioritized clonotypes shared across sample types (tumor, cfDNA, and PBMC), recurrent across patients, or detected at multiple tumor timepoints within individual patients. Among the selected sequences, 34 appeared in both tumor and either cfDNA or PBMC samples, 21 recurred in more than two patients, and 81 were detected across paired biopsy and resection samples within patients. These criteria were not mutually exclusive (Supplementary Table 1). We incorporated the resulting TCR panel into a 477-probe Xenium assay for *in situ* spatial transcriptomic profiling (Fig. 1c, Supplementary Table 2) and profiled paired biopsy and resection FFPE tumors from five HRNB patients, yielding 2,072,978 cells across 10 samples (Fig. 1d).

We first performed batch correction and integrated all 10 samples into a single dataset using Harmony^23^ and Leiden clustering^24^. We identified seven major cell types: neuroblast cells (n = 1,220,645), Schwann cells (n = 69,405), fibroblasts (n = 161,654), endothelial cells (n = 121,759), macrophages (n = 261,621), B cells (n = 59,432), and T cells (n = 178,462) (Fig. 1d,e). All seven major cell types were detected at both biopsy and resection in all patients (Fig. 1d, right)^14,25^, and enabling longitudinal comparison of tumor and immune composition within matched tumor pairs.

Changes in neuroblast and immune cell proportions varied substantially between patients (Fig. 2). We defined total immune representation as the combined proportions of macrophages, B cells, and T cells. Three patients (Patients 1-3) showed reductions in neuroblast proportion from biopsy to resection (0.83→0.45, 0.68→0.58, and 0.36→0.33), accompanied by increases in immune representation (0.19→0.26, 0.14→0.42, and 0.18→0.31). In contrast, two patients (Patients 4-5) showed stable or increased neuroblast proportions (0.39→0.49 and 0.32→0.34) alongside reductions in immune representation (0.39→0.16 and 0.41→0.17). These patterns did not segregate by treatment arm, as Patients 1-2 received standard of care alone (Arm A) whereas Patients 3-5 received standard of care plus 131I-MIBG (Arm B). Given the small cohort size, these compositional shifts were interpreted descriptively and were not subjected to formal statistical testing. Together, these findings indicate heterogeneous patient-specific remodeling of the HRNB tumor microenvironment following therapy.

**Figure 2.**
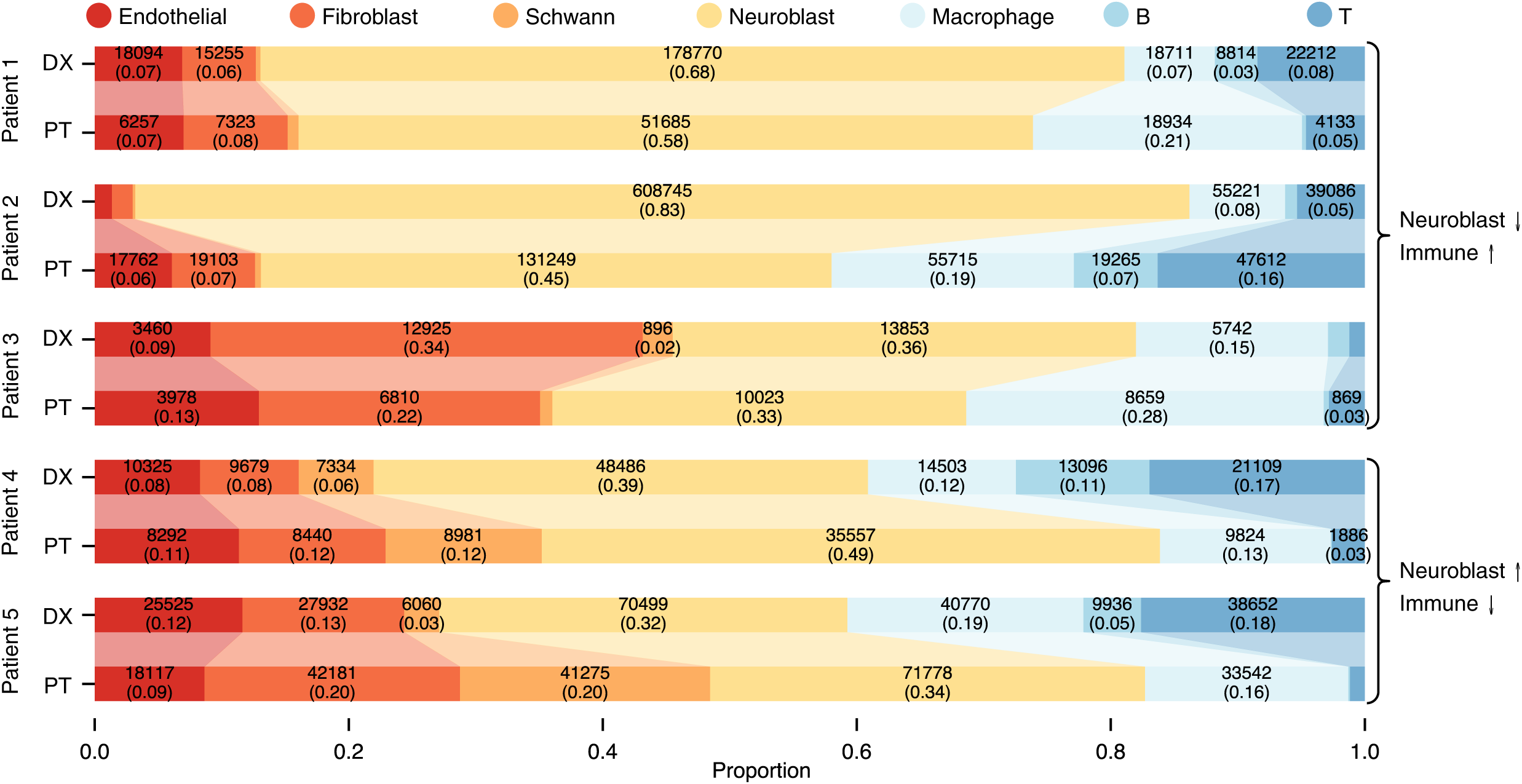
Tumor-immune compositional trajectories of primary HRNB tumors. Stacked horizontal bar plots of cell-type proportions across ten paired DX and PT samples (n = 5 patients). Alluvial ribbons connect the matched DX and PT bars for each patient. Bar annotations indicate per-segment cell counts and proportions.

### Cell-type co-localization analysis shows consistent tumor-immune segregation

To assess whether previously reported tumor-immune compartmentalization in neuroblastoma is recapitulated in our dataset, we quantified how different cell types are spatially organized relative to one another (Fig. 3). Specifically, we measured pairwise cell-type co-localization within a k-nearest neighbor spatial graph (k = 6), and compared observed frequencies to a null distribution generated by randomizing cell type labels while preserving cell positions. This approach tests whether specific cell types are found in proximity more or less often than expected by chance, providing a quantitative measure of spatial association or separation.

**Figure 3.**
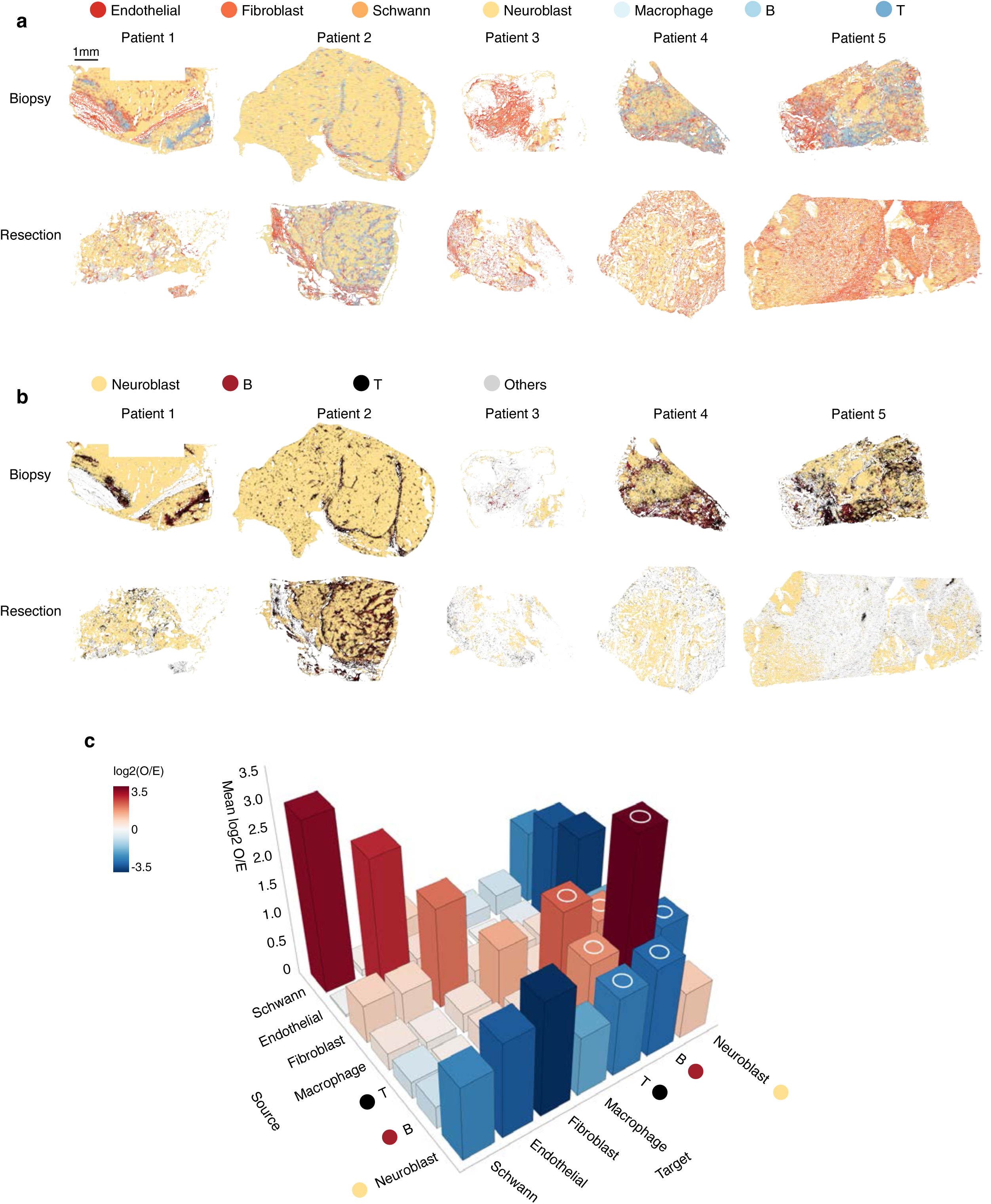
Spatial organization and tumor-immune remodeling of the HRNB tumor microenvironment. **a**, Spatial scatter plots of the ten FFPE specimens, showing the distribution of major cell types. Scale bar, 1 mm. **b**, Spatial scatter plots of the same specimens, showing neuroblasts in yellow, B cells in maroon, and T cells in black. Immune cells (blue + red) form clustered regions that are spatially distinct from neuroblast-enriched regions (yellow). **c**, Consensus pairwise spatial co-localization enrichment 3D barplot across all ten samples. Color indicates mean log2(observed/expected) interaction frequency, derived from a permutation-based null model (n = 5,000 permutations per sample). Circles denote reciprocal B-T, T-T, B-B, T-neuroblast and B-neuroblast interactions concordantly enriched or depleted across all ten samples.

Across all ten specimens, including paired biopsy and resection tumors, immune cells frequently occupied regions distinct from neuroblast-enriched areas (Fig. 3a,b). This pattern was supported by enrichment analysis, which showed consistent overrepresentation of T-T, B-B, and T-B proximities, and underrepresentation of interactions between adaptive immune cells (T and B cells) and neuroblasts (Fig. 3c; Extended Data Fig. 2). Together, these observations are consistent with prior reports of spatial compartmentalization in neuroblastoma^15^ and indicate that immune cells tend to localize near each other and away from tumor cells across patients and timepoints.

### T cell subtypes occupy distinct spatial niches, with γδT cells preferentially localized to neuroblast-rich regions

To further resolve T cell heterogeneity, we performed subclustering of T cells for all ten samples (Extended Data Fig. 3a-c). This analysis identified five major T cell subtypes: cytotoxic T cells, proliferating T cells, naive/central memory (CM) T cells, regulatory T cells (Tregs), and γδT cells (Extended Data Fig. 3b-c, Supplementary Table 3). Cytotoxic and proliferating T cell subsets exhibited the highest scores for exhaustion and tumor-infiltrating lymphocyte (TIL) dysfunction signatures^26,27^, whereas naive and central memory (CM) T cells showed the lowest scores (Extended Data Fig. 3d-f). Exhaustion and dysfunction signatures also trended higher after initial therapy across patients, although these changes did not reach statistical significance (Extended Data Fig. 3g,i). Across the five paired tumor cases, diagnosis-to-resection changes in these signature scores did not show a consistent pattern by treatment arm (two-sided Fisher’s exact test for direction of change: cytotoxicity, P=1.0; exhaustion and TIL dysfunction, not tested because all cases increased) (Extended Data Fig. 3g-i, Extended Data Fig. 3 source data).

Next, to assess whether specific T cell subtypes were able to bypass overall tumor-immune segregation and localize near neuroblasts, we performed a top-ten neighbor analysis, quantifying the cell type composition of the ten nearest neighbors for each T cell (Fig. 4). Across all samples, γδT cells and Tregs showed the highest proportion of neuroblast neighbors (Fig. 4a), a pattern that was consistent across biopsy and resection timepoints (Extended Data Fig. 4a). For γδT cells, neuroblasts comprised a mean proportion of 0.45 of the ten nearest neighbors (range 0.37-0.59) and were the most frequent neighboring cell type in all of the ten specimens analyzed (Extended Data Fig. 4a). Tregs showed a similar enrichment (mean 0.42; range 0.28-0.52). In contrast, naive/CM and proliferating T cells most frequently neighbored other T cells (mean 0.25 and 0.29, respectively), whereas cytotoxic T cells most commonly neighbored macrophages (mean 0.28) (Fig. 3a). These subtype-specific spatial patterns are consistent with the reported antitumor and phosphoantigen-sensing capacity of γδT cells in neuroblastoma^28,29^ and the enrichment of Tregs within immunosuppressive tumor niches^30^.

**Figure 4.**
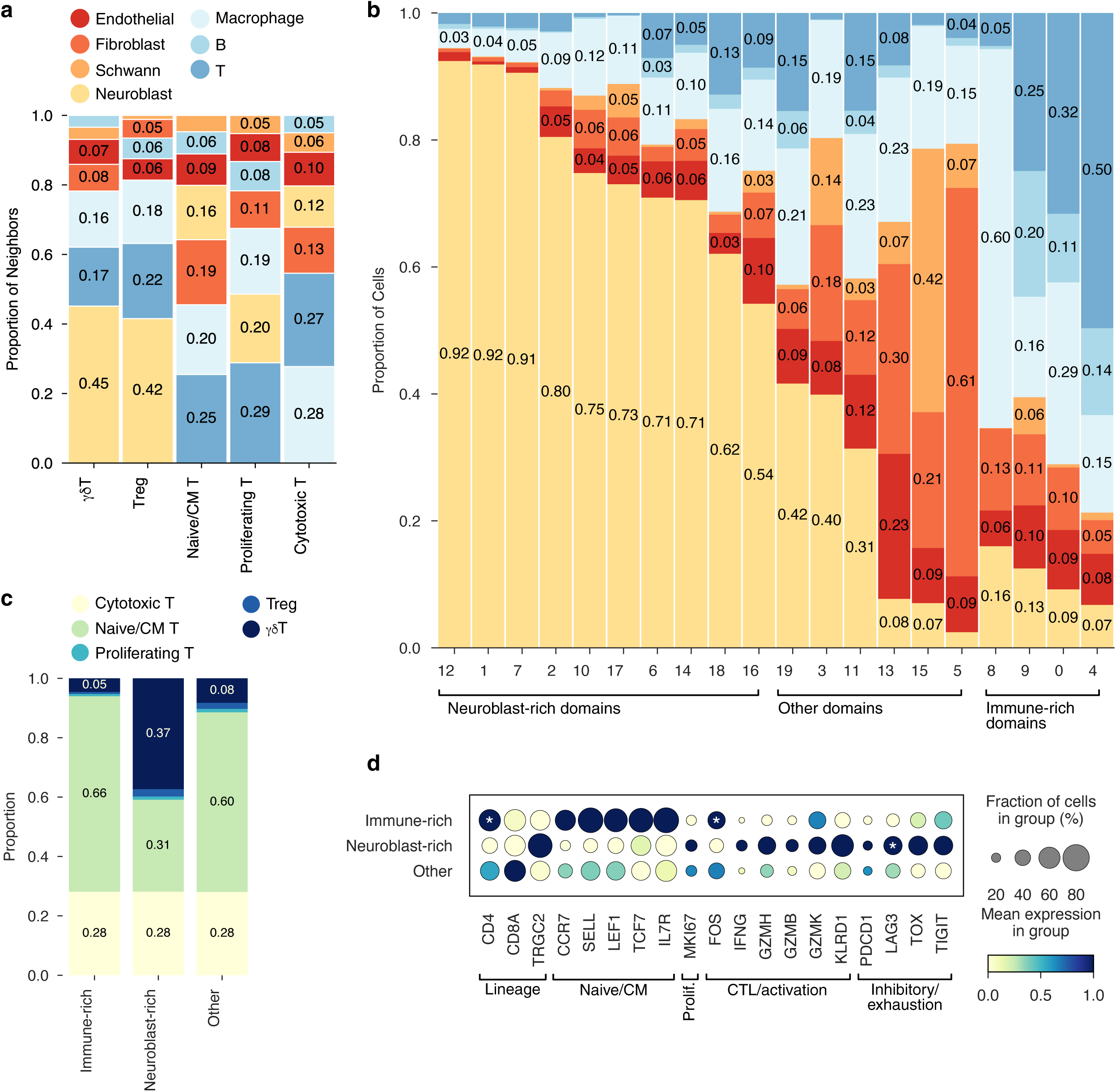
Top-10 proximity analysis, tissue domain architecture and spatially stratified T-cell transcriptional states. a, Stacked bar plot showing the mean composition of ten nearest cells for each T-cell subtype, aggregated across all ten samples with equal sample weighting. Bar segments colored by major cell type; labels indicate proportions ≥ 0.05. b, Stacked bar plot of cell-type composition across 20 tissue domains identified by spatial neighborhood clustering within an 80 µm radius. Domains are ordered by decreasing neuroblast proportion, with four immune-enriched domains at right. Segment labels indicate per-domain proportions. c, Stacked bar plot of T-cell subtype composition across three spatial domain categories (immune-rich, neuroblast-rich, other), shown as consensus (left) and per sample (right). Domains were classified as immune-rich (total immune cell proportion ≥ 0.5) or neuroblast-rich (neuroblast proportion ≥ 0.5); remaining domains were classified as other. d, Dot plot of T-cell marker expression across the three domain categories. Markers shown span lineage, naive/central memory, proliferating, cytotoxic, and inhibitory programs. Dot size, fraction expressing; color, standardized mean expression. *, FDR < 0.05.

We next examined broader tissue organization using Scimap^31^ to define compositionally distinct cellular domains across the ten specimens. Domain identification was performed at a fixed model resolution, yielding 20 domains with distinct cell type proportions (Fig. 3b). To facilitate interpretation, we operationally defined domains containing at least 50% neuroblasts as neuroblast-rich, and domains containing at least 50% immune cells (macrophages, T cells, and B cells) as immune-rich. Four domains met criteria for immune-rich organization, with macrophages predominating in domain 8 (proportion 0.60) and T cells enriched in domains 9, 0, and 4 (proportions 0.25, 0.32, and 0.50, respectively).

After grouping domains into Immune-rich, Neuroblast-rich, and Other categories, we assessed T-cell subtype distribution across spatial contexts (Fig. 4c). Cytotoxic T-cell proportions did not differ across domains (Friedman test, BH-FDR P = 0.285). In contrast, Naive/CM T-cell proportions varied by domain (Friedman test, BH-FDR P = 0.038), with lower representation in neuroblast-rich versus other domains (paired Wilcoxon, BH-FDR P = 0.029). Treg proportions also differed across domains (Friedman test, BH-FDR P = 0.0038), with enrichment in neuroblast-rich domains relative to immune-rich and other domains (paired Wilcoxon, BH-FDR P = 0.029 for both contrasts). γδT cell proportions showed strong domain dependence (Friedman test, BH-FDR P=0.0038) and were enriched in neuroblast-rich versus other domains (paired Wilcoxon, BH-FDR P = 0.029). This neuroblast-rich enrichment of γδT cells was consistently observed across samples containing γδT cells (Extended Data Fig. 4b).

To determine whether T cell states differed by spatial context, we compared domain-stratified T cell expression across immune-rich and neuroblast-rich regions (Fig. 4d). Paired pseudobulk testing across samples (Wilcoxon signed-rank, BH-FDR) identified higher *LAG3* and lower *CD4* and *FOS* expression in neuroblast-rich versus immune-rich domains (all FDR = 0.0247; n = 10 paired samples)^27,32^. Other lineage, naive/CM, proliferation, cytotoxic, and inhibitory markers showed directional trends across domains but were not significant after multiple-testing correction.

Together, these findings indicate that γδT cells are preferentially positioned within neuroblast-rich territories and that neuroblast-rich domains are associated with higher *LAG3* and lower *CD4* and *FOS* expression in the T cell compartment relative to immune-rich domains^27^. These data align with a recent report identifying γδT cells as prominent antitumor effectors in HRNB^33^, and support a model in which γδT cells constitute a spatially privileged T cell population within neuroblast-rich regions of HRNB.

### Spatial TCR profiling resolves patient-specific clonotypes

Having established that T cell subtypes occupy distinct spatial niches, we next asked whether individual TCR clonotypes could be localized within the same tissue sections. Because FFPE-based *in situ* detection may capture off-target or low-confidence TCR probe signals, we applied stringent *post hoc* filtering to the Xenium data (Extended Data Fig. 5). To retain high-confidence clonotype-specific signals, each candidate CDR3 probe had to meet three criteria: 1) show at least two-fold enrichment in T cells relative to non-T cells in the Xenium specimen from the same patient in which that clonotype was identified by bulk CapTCR-seq; 2) be detected in at least 10 T cells; and 3) show concordant expression with chain-specific markers (CD3E with CD4 or CD8A for TCRα/β probes; CD3E with TRGC2 for TCRγ/δ probes).

Of the 100 candidate probes, 24 met all criteria, including 6 TCRγ, 3 TCRδ, 8 TCRα and 7 TCRβ probes (Fig. 5a). These 24 high-confidence TCR sequences were distributed across seven of the ten tumor specimens, defining a spatially validated set for clonotype-level analysis. We next identified paired clonotypes at single-cell resolution by requiring co-detection of TCRα and TCRβ CDR3 probes within the same cell. This approach recovered four paired αβ clonotypes, corresponding to eight of the 24 high-confidence probes. The remaining 16 probes were retained as unpaired clonotype detections, yielding 20 total spatially resolved clonotypes. We did not recover paired γδ clonotypes, likely reflecting the limited number of TCRδ sequences captured in the original bulk repertoire.

**Figure 5.**
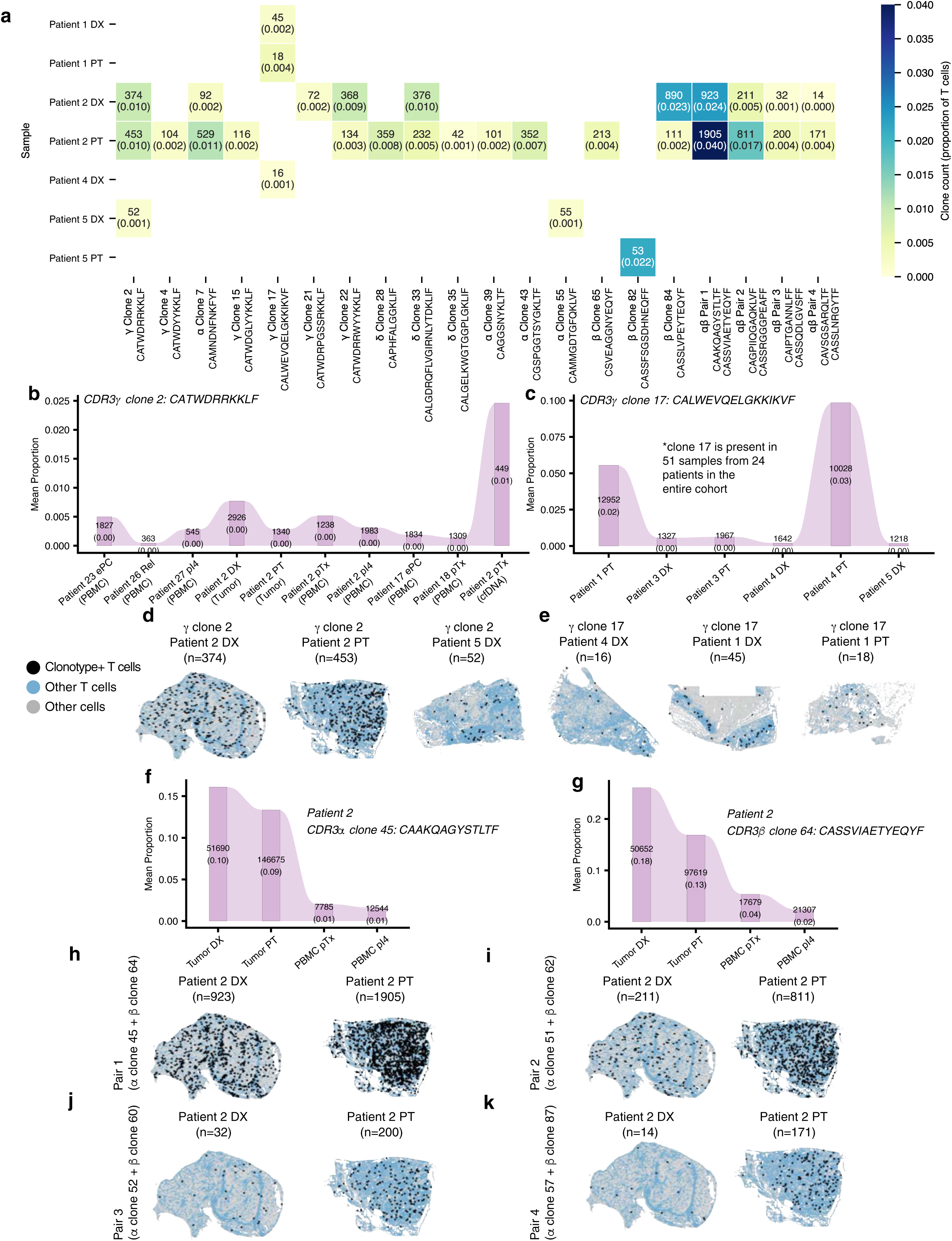
Spatial TCR clonotype profiling successfully tracks 24 TCRs across paired tumor biopsies and resections. **a**, Heatmap of the 24 validated CDR3 probes (represented as 20 unique clonotype entities after collapsing αβ-paired chains) across all ten FFPE samples. Rows indicate samples ordered as DX and PT pairs per patient; columns represent individual clonotype entities grouped by TCR chain class (γ, δ, unpaired α, unpaired β, αβ pairs). Color encodes the proportion of T cells that were clonotype-positive; cell annotations show absolute count and proportion. Cells are masked where the probe was not targeted in that patient. **b**, Alluvial clonotype tracking plot for TCRγ clone 2 (CATWDRRKKLF) across tumor, PBMC and cfDNA from six patients (899532, 904847, 907444, Patient 2, 909615 and 910227; n = 10 tumor samples). **c**, Alluvial clonotype tracking plot for TCRγ clone 17 (CALWEVQELGKKIKVF) across paired tumor DX and PT samples from four patients (Patient 1, Patient 3, Patient 4, and Patient 5; n = 6 tumor samples). **d**, Spatial scatter plots of γ clone 2-positive T cells (maroon) in Patient 2 DX, Patient 2 PT, and Patient 5 DX samples. All T cells are shown in blue; non-T cells in gray. Clonotype-positive cells were defined by the co-detection of the CDR3-targeting probe together with TRGC2, and CD3E within the same cell. **e**, Spatial scatter plots of γ clone 17-positive T cells in Patient 4 DX, Patient 1 DX, and Patient 1 PT samples. **f**, Alluvial clonotype tracking plot for TCRα clone 45 (CAAKQAGYSTLTF) across PBMC (pTx and pI4) and tumor (DX and PT) samples from Patient 2. Band width reflects clonotype proportion; labels indicate read count and within-sample proportion. **g**, Alluvial clonotype tracking plot for TCRβ clone 64 (CASSVIAETYEQYF) across the same samples from Patient 2, as in **f**. **h**-**k**, Spatial scatter plots of αβ clonotype pair-positive CD8+ T cells in Patient 2 DX and PT samples. Clonotype-positive cells required co-detection of both paired CDR3 probes together with CD8A and CD3E within the same cell: αβ Pair 1 (**h**, α clone 45 + β clone 64), αβ Pair 2 (**i**, α clone 52 + β clone 60), αβ Pair 3 (**j**, α clone 51 + β clone 62) and αβ Pair 4 (**k**, α clone 57 + β clone 87).

Among the 20 spatially resolved clonotypes, 10 were detected at both biopsy and resection, and two TCRγ clonotypes were shared across patients (Fig. 5a). Clonotype recovery varied across specimens, with 16 of 20 clonotypes detected in PBBCHF. For clonotypes observed at both timepoints, we classified biopsy-to-resection dynamics using clonotype fraction ratios consistent with Immunarch^34^ repertoire analysis thresholds (expanded, >1.10; contracted, <0.90; stable, 0.90-1.10). This identified six expanding, three contracting and one stable clonotype, indicating heterogeneous longitudinal clonal dynamics across paired HRNB tumors.

### Spatial context associates with transcriptional state within shared γδ TCR clonotypes

To test whether spatial location within the tumor microenvironment was associated with transcriptional state within the same TCR-defined clonotype, we integrated clonotype-resolved TCR probe calls with scimap-derived immune-rich, neuroblast-rich, and other domain annotations (Fig. 5; Extended Data Fig. 6, Extended Data Fig. 7a).

We first examined two shared γδ clonotypes to capture potentially public or conserved tumor-associated responses. Vγ8-associated γ clone 2 (CATWDRRKKLF) was detected across multiple compartments and patients, including Patient 2 (biopsy and resection, tumor, PBMC and cfDNA), Patient 5 (biopsy), and five additional individuals with blood samples only, and remained proportionally stable across timepoints in Patient 2 (0.010) (Fig 5b, Extended Data Fig. 6a). Within immune-rich domains, clone 2-positive T cells expressed higher levels of *TCF7*, *LEF1*, *SELL* and *IL7R*^35^ compared to other T cells, consistent with a less differentiated transcriptional program (Extended Fig 7a). In contrast, clone 2-positive cells in neuroblast-rich domains showed increased expression of cytotoxic effector genes (*IFNG*, *KLRD1*)^36,37^ and dysfunction-associated markers (*TOX*, *LAG3*)^27,38,39^, although these trends did not reach statistical significance (Extended Fig 7a).

A second shared clonotype, Vγ9-associated γ clone 17 (CALWEVQELGKKIKVF), was detected in Patient 4 (biopsy) and Patient 1 (biopsy and resection) and increased proportionally in Patient 1 from 0.002 to 0.004 (Fig. 5c, Extended Data Fig. 6b). Domain-stratified comparisons showed a similar directional pattern to clone 2, with immune-rich domains associated with higher expression of naive/CM-associated genes and neuroblast-rich domains associated with higher *TOX* and *LAG3*, although statistical significance was limited by clone abundance (Extended Fig. 7a). Clone 17 was detected in 51 samples across 24 patients in the entire cohort, and was consistent with a circulating public Vγ9-Vδ2-associated γδ clonotype^40,41^. While this clonotype has been widely reported in healthy blood as a public clone, the exact antigen recognized by this TCR has yet to be reported.

Together with the enrichment of γδ T cells in neuroblast-rich regions, these findings support a model in which shared γδ clonotypes exhibit spatially associated shifts in transcriptional state across tumor regions. Detection of clone 2 and clone 17 across patients did not align with treatment-arm assignment, suggesting that these shared γδ clonotypes were unlikely to represent ^131^I-MIBG treatment-associated antigen responses.

### Paired αβ clonotypes revealed domain-dependent transcriptional divergence and neoantigen-associated programs

Because bulk sequencing did not resolve αβ chain pairing, we leveraged Xenium co-detection to identify paired αβ clonotypes at single-cell resolution. Four CD8+ αβ clonotype pairs, prioritized from bulk immunosequencing, were validated by *in situ* co-detection of α and β CDR3 probes within the same cells (Fig. 5f-k, Extended Data Fig. 7c-f). In contrast, no γδ clonotypes were successfully paired, likely reflecting lower transcript abundance and/or probe coverage for γ and δ chains. Despite contraction of bulk TCR diversity in blood and tumor (Extended Data Fig. 1), all four αβ pairs were expanded from biopsy to resection in Patient 2, with distinct magnitudes (Fig. 5a,h-k).

Across clonotype pairs, spatial context is consistently tracked with transcriptional state. Cells from the same clonotype located in immune-rich domains expressed higher *LEF1* and *SELL*, whereas those in neuroblast-rich domains showed higher *THEMIS*^42^, *IFNG* and *TOX* (Extended Fig. 7a). Pair-level analysis provided additional resolution: pair 1 (Fig. 5f-h, Extended Fig. 6c) exhibited high *THEMIS* and TCF7 with low *LAG3* in neuroblast-rich domains, consistent with a precursor-like dysfunctional state^43,44^ (Extended Fig. 7a). Pair 2 (Fig. 5i, Extended Fig. 6d) recapitulated this structure, with immune-rich enrichment of *TCF7*, *LEF1*, *SELL* and *IL7R*^35^ and neuroblast-rich enrichment of *GZMH*^45^ (Extended Fig. 7a). Pairs 3 and 4 showed similar directional trends but did not meet minimum cell thresholds for formal statistical testing (Fig 5j,k, Extended Fig. 6e,f, Extended Fig. 7a).

To assess whether paired αβ clonotypes exhibit features consistent with tumor antigen engagement, we scored clonotype-positive and clonotype-negative CD8+ T cells using four published neoantigen-reactive CD8+ TIL gene signatures^46–50^, restricted to genes present on the Xenium panel (Extended Data Fig. 7b; Supplementary Table 4). Pair 2 showed the most consistent enrichment, with all four signatures higher in clonotype-positive cells in specimens from diagnostic biopsy and primary tumor resection (FDR < 0.05; Caushi, Oliveira, Lowery and Hanada, each q < 0.001 at one or both timepoints). Pair 3 was enriched for Caushi, Oliveira and Lowery at both timepoints and for Hanada at resection only (FDR < 0.05). Pair 1 showed mixed results: Oliveira, Lowery and Hanada scores were higher in clonotype-positive cells (FDR < 0.05), whereas the Caushi score was lower at both timepoints. Together, these findings suggest that some paired αβ clonotypes display transcriptional features associated with neoantigen-reactive CD8+ T cells, with variable concordance across signatures.

Collectively, these results show that spatial context is associated with transcriptional state even within identical TCR-defined clonotypes. Additionally, all four paired αβ clonotypes scored higher for neoantigen-reactive TCR markers and expanded post-therapy within the tumor. Coupled with the decrease in TCR diversity following initial therapy observed in both PBMCs and tumors (Extended Data Fig. 1a-f), this expansion points to antigen-driven clonal expansion.

## Discussion

Here, we integrated longitudinal bulk TCR sequencing with Xenium-based spatial transcriptomic profiling to characterize how the immune microenvironment of HRNB relates to T cell state at single-clonotype resolution (Fig. 6). Across paired diagnostic biopsy and primary tumor resection specimens, we found a consistent spatial compartmentalization characterized by segregation between tumor and immune compartments and co-localization among immune cells. Within this global structure, however, specific T cell subsets occupied distinct spatial niches. γδT cells and Tregs were preferentially positioned adjacent to neuroblasts, and γδT cells were enriched within neuroblast-rich domains. These observations indicate that tumor-immune segregation is not uniform, but instead coexists with localized interfaces where specific immune populations engage tumor cells.

**Figure 6.**
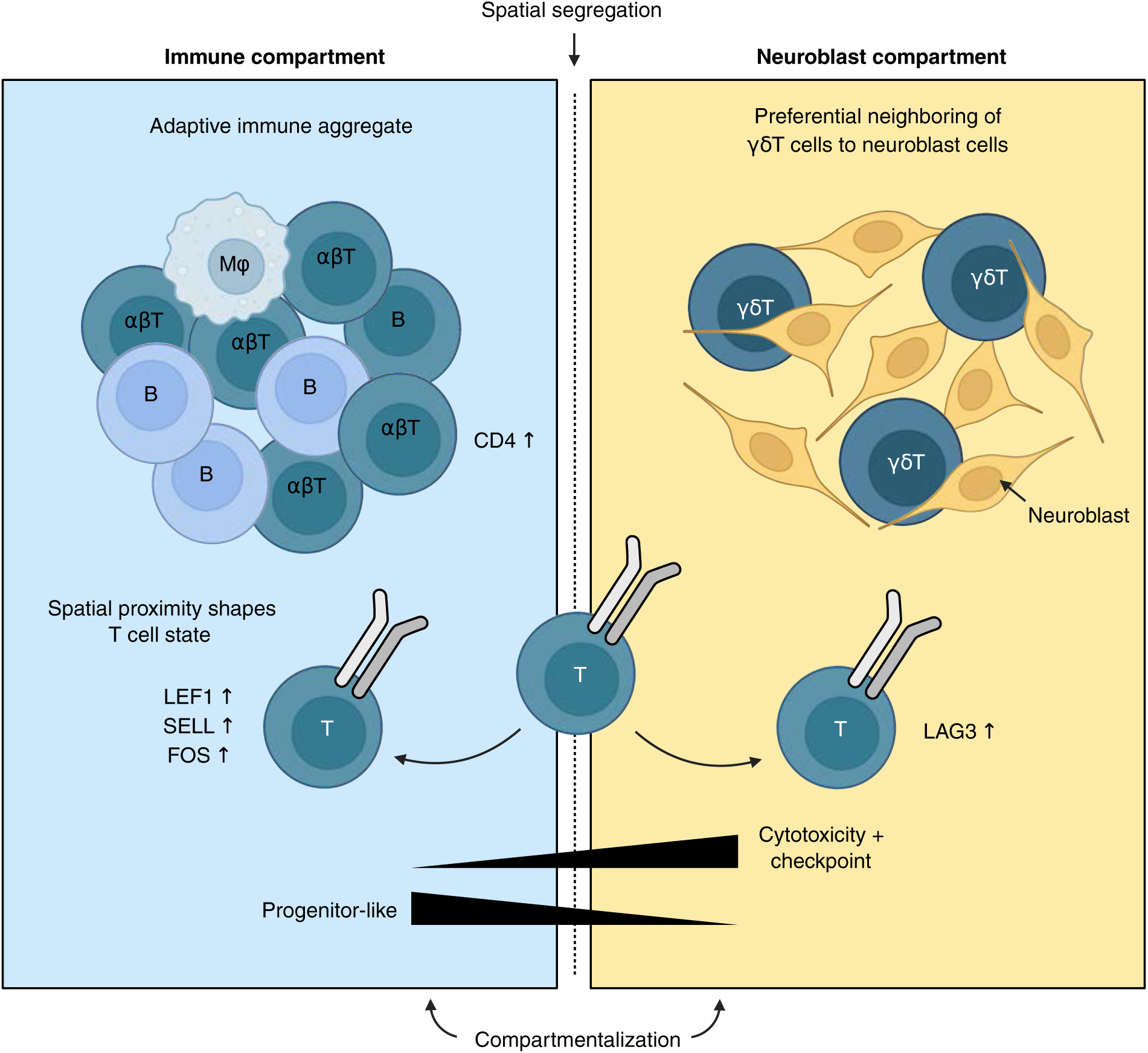
Concept diagram of spatial immune organization and clonotype-state bifurcation in HRNB. The HRNB microenvironment is divided into spatially segregated immune-rich and neuroblast-rich compartments. The immune compartment contains an adaptive aggregate with treatment-associated upregulation of *CCL4*-*CCR5* and *CCL3*-*CCR4*. The neuroblast compartment is characterized by preferential positioning of γδ T cells. The same αβ T cell clonotype exhibits a progenitor-like program (*LEF1*, *SELL*, *FOS*) in immune-rich regions and a checkpoint-associated program (*LAG3*) at the tumor interface, represented by opposing spatial gradients. Created with BioRender.com.

A key finding of this study is that spatial context is associated with transcriptional state even within identical TCR-defined clonotypes. Across both shared γ/δ clonotypes and paired CD8+ αβ clonotypes, immune-rich domains preferentially supported less differentiated programs marked by *TCF7*, *LEF1*, *SELL* and *IL7R*, whereas neuroblast-rich domains were associated with cytotoxic and dysfunction-related signatures, including *IFNG*, granzymes, *KLRD1*, *TOX* and *LAG3*. These results suggest that T cell state is not solely determined by clonotype identity, but is shaped by local microenvironmental context. More broadly, our findings reframe HRNB from an immune “cold” tumor to a spatially immune-structured ecosystem organized into immune-permissive and tumor-proximal zones that differentially support progenitor-like versus cytotoxic/checkpoint-associated programs. This spatial heterogeneity is consistent with recent reports in adult cancers, where T cell states vary across tissues and maintain a more progenitor-like state in immune-rich niches^19,51^.

The longitudinal bulk TCR analysis further supports a dynamic relationship between systemic immunity, circulating repertoires and the tumor microenvironment in HRNB. Across matched compartments, cfDNA repertoires showed greater overlap with tumor-associated clonotypes than PBMC repertoires. Although cfDNA contained relatively few detectable clonotypes overall, these findings suggest that the circulating cfTCR repertoire may preferentially capture tumor-proximal immune features not readily observed in peripheral blood alone. This observation aligns with recent work demonstrating that tumor microenvironment-associated spatial ecotypes can be inferred from plasma cfDNA across multiple cancer types^52^, supporting the broader concept that cfDNA contains biologically informative signals derived from tumor-associated cellular ecosystems.

Neuroblastoma is characterized by altered metabolic programs, including increased pyruvate flux and activation of the mevalonate pathway, which can elevate intracellular phosphoantigen levels^53^. These metabolites are known ligands for Vγ9Vδ2 TCRs and can promote γδ T cell activation. The enrichment of γδ T cells within neuroblast-rich regions, together with the presence of a shared Vγ9-associated clonotype (clone 17), is consistent with local metabolite-driven activation in the tumor microenvironment^53,54^. However, antigen specificity and metabolite availability were not directly measured in this study, and these interpretations remain speculative. Future studies integrating spatial metabolomics or functional perturbation will be required to test this hypothesis.

This study has several limitations that motivate the next phase of work. First, FFPE preservation likely reduces recovery of cytosolic RNA, limiting sensitivity for CDR3 detection and low-abundance transcripts. Clonotype detection was therefore uneven across samples. Second, bulk-informed probe selection introduces bias toward higher-frequency clonotypes and limits the number of paired αβ clonotypes that can be resolved. Third, the cohort size is small, and clone-positive cell counts are sparse within domains, limiting statistical power for domain-stratified and pair-level analyses. Fourth, the targeted Xenium panel samples only a subset of the transcriptome, constraining pathway-level interpretation and overlap with published gene signatures. Finally, survival data are not yet available for ANBL1531, and although ^131^I-MIBG arm assignment was available for the five-patient spatial cohort, the small sample size, limited clinical outcome data, and possibility of heterogeneous or limited treatment effect precluded formal arm-specific or outcome-correlative analyses of spatial features. Altogether, these limitations motivate ongoing efforts toward whole-transcriptome spatial profiling with unbiased TCR capture.

Despite these constraints, this study provides a framework for linking T cell clonotype, spatial context and transcriptional state in HRNB. We identify neuroblast-rich domains as a key spatial axis associated with T cell differentiation and dysfunction, highlight γδ T cells as a prominent tumor-adjacent population, and suggest that spatial organization may influence both T cell state and antigen-associated activity. These findings support the idea that therapeutic strategies in HRNB may benefit from considering spatial context, including approaches that target tumor-immune interfaces, modulate checkpoint-associated states, or leverage γδ T cell responses.

## Methods

### Patient cohorts and study design

This study was conducted using specimens collected via a Children’s Oncology Group (COG) (U10CA180886, U10CA180899, cIRB OMB#0925-0753, IND# 134379) trial, and was approved by the Children’s Hospital of Philadelphia (CHOP) committee for protection of human subjects, and the University Health Network (UHN) Research Ethics Committee (CAPCR-ID: 22-5027.3). Specimens were obtained through COG ANBL1531. Written informed consent was obtained from parents or legal guardians in accordance with the Declaration of Helsinki.

Patients were newly diagnosed with high-risk neuroblastoma who met eligibility were enrolled on Arms A or B of the ANBL1531 clinical trial (NCT03126916). Patients on both arms received standard-of-care therapy, with Arm B patients additionally receiving a single dose of ^131^I-metaiodobenzylguanidine (^131^I-MIBG) prior to induction cycle 4. Clinical sampling was anchored to predefined trial milestones. For tumor tissue, FFPE specimens were collected at the time of diagnostic biopsy and at post-chemotherapy surgical resection, which typically occurred after cycle 4. For peripheral blood-derived specimens (cfDNA and PBMCs), longitudinal milestones included diagnosis, pre-induction cycles 2, 3 and 4, 72 hours post-^131^I-MIBG treatment, pre-surgical resection, end-of-induction, end-of-post-consolidation, progression, and relapse.

The current study included five patients with paired FFPE biopsy and resection tumors (n = 10 tumor specimens), along with additional peripheral blood-derived specimens including cfDNA (n = 14) and PBMCs (n = 84), collected longitudinally where available. In total, 15 specimens from the paired tumor cohort (including matched cfDNA and PBMC samples when available) and an additional set of blood-derived samples were subjected to bulk TCR sequencing (CapTCR-seq), enabling longitudinal tracking of the TCR repertoire across clinical timepoints. All ten FFPE tumor sections were further profiled using Xenium spatial *in situ* gene expression with a combined transcriptomic and targeted CDR3 probe panel.

Specimen processing and inter-site transfer followed standard biobanking workflows. Whole blood for cfDNA was collected in Streck tubes at COG study sites and shipped to BPC for plasma isolation; plasma was subsequently transferred through Dana-Farber Cancer Institute and the Broad Institute before bulk TCR sequencing at UHN. Whole blood for PBMC isolation was collected in EDTA tubes, processed at CHOP, and shipped to UHN. PBMCs were cryopreserved in FBS supplemented with 10% DMSO (1 mL per vial). For cfDNA, bulk sequencing was performed using 200 ng input DNA (typical elution volume 29 μL). Xenium imaging and initial data generation from FFPE tumor sections were performed at CHOP, whereas tumor-derived DNA for CapTCR-seq was shipped to UHN for bulk sequencing; processed data were transferred to UHN for downstream integration and analysis.

### Sample processing, capture library preparation and CapTCR-seq

Hybrid-capture targeted immunosequencing (CapTCR-seq), an in-house method previously described^20^, was performed on FFPE tumor genomic DNA, PBMC genomic DNA and plasma-derived cell-free DNA (cfDNA) to profile TCRα, TCRβ, TCRγ and TCRδ repertoires. Capture baits targeted V and J gene segments spanning CDR3-containing regions across TRAV/TRAJ, TRBV/TRBJ, TRGV/TRGJ and TRDV/TRDJ loci. Genomic DNA from FFPE tumor tissue and PBMCs was extracted using the AllPrep DNA/RNA/miRNA Universal Kit (QIAGEN), and cfDNA was isolated using the QIAamp Circulating Nucleic Acids Kit (QIAGEN). DNA yield was quantified using Qubit, and fragment size distributions were assessed using TapeStation or Bioanalyzer.

Tumor and PBMC genomic DNA were fragmented by ultrasonication (Covaris LE220) to a target library insert size of ∼250 bp, whereas cfDNA was used without additional fragmentation. Sequencing libraries were prepared using KAPA HyperPrep (Roche) with IDT Duplex-Seq adapters and unique dual indices (UDIs) following the manufacturers’ protocols. Input DNA amounts were standardized by specimen type (FFPE tumor DNA, 500 ng; PBMC genomic DNA, 150 ng; cfDNA, 20 ng).

Target enrichment was performed using an IDT xGen™ Custom Hyb Panel (Accel design) and the IDT xGen™ hybridization capture of DNA libraries protocol with custom V/J probe sets. Following hybridization, captured libraries were recovered using streptavidin bead-based pull-down and washed under stringent conditions. To reduce off-target background and enrich for CDR3-containing fragments, libraries underwent sequential enrichment steps, including depletion hybridization followed by reverse capture, and subsequent V-probe hybridization as described in the CapTCR-seq method^15^. Hybridization was performed at 50°C with overnight incubation following the published thermocycler program.

Sequencing was performed in three staged rounds to confirm capture performance before deep profiling. First, pre-capture libraries were sequenced at shallow depth (10^3^-10^6^ read pairs) to evaluate library complexity and barcode representation. Second, post-capture QC libraries were sequenced on an Illumina MiSeq to confirm on-target enrichment. Third, post-capture deep sequencing was performed on an Illumina NovaSeq 6000 using a 300-cycle kit (2 × 150 bp).

### CapTCR-seq read processing and TCR repertoire analysis

Sequencing reads were processed using MiXCR v4.6.0 (preset: hsa) for alignment, V(D)J assignment and clonotype assembly^55,56^. Because CapTCR-seq enriches short CDR3-containing fragments rather than full-length receptors, clonotypes were defined by unique CDR3 amino-acid sequences within each chain (TCRα, TCRβ, TCRγ and TCRδ). Only productive, in-frame clonotypes were retained for downstream analyses.

Repertoire analyses were performed in R (v4.4.2)^57^ using Immunarch (v0.10.3)^34^ on MiXCR outputs. Repertoire metrics were computed for all chains, but only the TRB chain was shown to avoid redundancy. PBMC metrics were evaluated longitudinally across diagnosis, pre-induction cycle 4, pre-surgical resection and end-of-post-consolidation, whereas tumor metrics were compared between matched diagnostic biopsy and post-therapy resection specimens. Shannon diversity was quantified using the Hill diversity framework (q = 1) as implemented in Immunarch, and repertoire volume was defined as the number of unique clonotypes per sample. Clone-size distributions were computed from within-sample relative clonotype abundance using bins small (≤0.0001), medium (≤0.001), large (≤0.01) and hyperexpanded (>0.01). For longitudinal PBMC metrics, we fit linear mixed-effects models using lme4 and performed pairwise timepoint contrasts using estimated marginal means in emmeans with Holm correction. Clonotype tracking across samples was performed using Immunarch trackClonotypes keyed to CDR3 amino acid identifiers.

### Xenium CDR3 probe selection and *post hoc* specificity filtering

To enable spatial tracking of T cell clonotypes, we designed a custom set of 100 add-on CDR3-targeting probes for Xenium based on bulk CapTCR-seq repertoires. Candidate CDR3 amino-acid sequences were selected from tumor, PBMC and cfDNA repertoires, with priority given to clonotypes detected in tumor over blood-only clonotypes. Candidate targets were then submitted to the 10x Genomics Xenium Panel Designer for probe design and manufacturability assessment, and finalized for synthesis as custom Xenium CDR3 probes. Because bulk sequencing does not provide αβ pairing, candidate TCRα and TCRβ sequences were additionally prioritized when their abundance patterns were highly concordant across matched tumor and peripheral compartments, nominating putative αβ pairs for subsequent single-cell co-detection.

A total of 100 CDR3 probes were selected across five patients and two timepoints, with probe allocation proportional to the number and diversity of candidate tumor-associated clonotypes per specimen. Allocation by specimen was: Patient 2 DX (22), Patient 2 PT (35), Patient 3 DX (22), Patient 3 PT (20), Patient 1 DX (7), Patient 1 PT (13), Patient 4 DX (27), Patient 4 PT (12), Patient 5 DX (25) and Patient 5 PT (8). Probes were distributed across TCR chains as follows: 44 TRB, 21 TRA, 23 TRG and 12 TRD. Probe sequences were submitted for custom Xenium synthesis and incorporated into the Xenium panel.

To identify clonotype-specific probes suitable for downstream spatial analyses, we evaluated probe performance *post hoc* using Xenium single-cell count matrices and cell-type annotations. Filtering was performed within the intended patient specimen(s) for each probe using three sequential criteria.

**Criterion 1:** T cell enrichment (mean expression). For each probe, we computed the mean raw probe counts per cell within T cells and within non-T cells. A probe was required to show at least a two-fold higher mean raw expression in T cells than in non-T cells in its intended specimen. T cells were defined by the Xenium-derived cell-type annotation used throughout the study. Probes failing this enrichment threshold were excluded.

**Criterion 2:** minimum detection frequency (binary detection). Probes passing criterion 1 were required to be detected in at least 10 T cells within the intended specimen, where detection was defined as a nonzero raw probe count (binary presence/absence per cell).

**Criterion 3:** chain concordance (normalized expression). Probes passing criteria 1-2 were required to show concordant expression with expected chain-associated marker patterns using normalized gene expression values. Normalization was performed by library-size normalization to a target sum of 1e^4^ counts per cell using Scanpy (sc.pp.normalize_total, target_sum=1e^4^), followed by log-transformation. For TRA/TRB probes, probe-positive cells were required to align with canonical T cell marker expression (CD3E) and either CD4 or CD8A. For TRG/TRD probes, probe-positive cells were required to align with CD3E and TRGC2. Probes inconsistent with the expected chain context were excluded.

Following filtering, the retained high-confidence probe set comprised 24 CDR3 probes (8 TRA, 7 TRB, 6 TRG and 3 TRD), which were used for downstream clonotype-resolved spatial analyses.

### Gene probe selection for the Xenium custom panel

In addition to the 100 customized CDR3 probes, we designed a customized 360-gene panel to detect the major malignant and immune populations and sub-populations in HRNB, based on our recent single-cell profiling^14^. Specifically, we selected the top 10-20 upregulated genes for each cell type, malignant state and macrophage subset. We also added an additional 30 genes based on literatures to better characterize T cell subsets, which were not the primary focus of our previous study. Beyond the CDR3 and gene probes, we designed 17 probes to detect known mutations in HRNB, including single-nucleotide variants in ALK, MYCN, BRAF, FGFR1 and ATRX fusion, collected from the COSMIC (https://www.sanger.ac.uk/tool/cosmic/) and PeCan (https://pecan.stjude.cloud/) databases.

### Xenium tissue processing, imaging and segmentation

The ten FFPE tumor sections were processed using the Xenium *In Situ* Gene Expression workflow (Xenium chemistry v1) according to the manufacturer’s protocol. Imaging and decoding were performed on the Xenium Analyzer (instrument software v4.0.1.4)^58^, and per-transcript count matrices and segmentation masks were exported for downstream analysis. Run performance was assessed using Xenium summary outputs (metrics_summary.csv), including decoding yield, negative control rates, estimated false-positive burden and segmentation summary metrics.

Cell segmentation was performed using 10x Genomics Xenium Onboard Analysis (v4.0.1.0)^58^ with the Multimodal Cell Segmentation algorithm and the standard stain set xenium_cell_segmentation_stains_v1 (DAPI; 18S interior RNA stain; boundary stain cocktail ATP1A1/CD45/E-cadherin; and αSMA/vimentin). Segmentation quality was evaluated using the fraction of transcripts assigned within cell masks, the fraction of empty cells and segmentation method attribution metrics. Across sections, segmented_cell_stain_frac exceeded 0.88 and nucleus-expansion segmentation remained a minority component (segmented_cell_nuc_expansion_frac ≤ 0.112), consistent with segmentation primarily supported by stain-informed signals.

### Xenium data processing, integration, and annotation

Spatial transcriptomic data were analyzed in Python using Scanpy^59^, Squidpy^60^ and SpatialData^61^. Cells were additionally visualized in Xenium Explorer (v4.1)^58^. TCR clone probes were excluded prior to this analysis. Negative control probes and codeword fractions were assessed as quality metrics. Cells and genes were filtered using sc.pp.filter_cells (min_counts = 20) and sc.pp.filter_genes (min_cells = 30), followed by normalization (target_sum = 1×10⁴) and log transformation. Integration across the ten tumor samples was performed using Harmony^23^ on sample labels, followed by neighbor-graph reconstruction (n_neighbors = 15) on

Harmony-corrected PCA^61^ embeddings (n_comps = 20), UMAP^63^ visualization, and Leiden^24^ clustering (resolution = 0.8). One low-quality cluster was identified by QC metric inspection and removed prior to cell type annotation. Marker genes were identified using Wilcoxon rank-sum tests, and cell identities were assigned by marker-guided annotation based on the targeted gene panel. T cells were subsequently subclustered using resolution = 0.5, yielding five refined T subtype annotations.

### Compositional analysis and longitudinal comparisons

Cell-type proportions were computed per sample and timepoint at the broad (seven major compartments) level. Longitudinal shifts in neuroblast and total immune-cell proportions were quantified as paired (resection - biopsy) deltas per patient.

Gene expression signature scores for cytotoxicity, exhaustion, and TIL dysfunction were computed on T cells using sc.tl.score_genes with gene sets derived from published signatures^26,27^, restricted to genes present in the Xenium targeted panel. Pseudobulk mean scores were derived per sample by averaging single-cell scores across all T cells in that sample (minimum 10 cells required). Longitudinal differences in pseudobulk scores between biopsy and resection were assessed using two-sided paired t-tests (scipy.stats.ttest_rel), with Wilcoxon signed-rank tests performed as a robustness check. P values from the three signature comparisons were jointly corrected using BH-FDR (statsmodels.stats.multitest.fdrcorrection).

UMAP embeddings for each lineage compartment were overlaid with per-timepoint cell density contours estimated by Gaussian kernel density estimation (scipy.stats.gaussian_kde)^64^ on a 200×200 grid, with contour levels defined at six evenly spaced intervals spanning the global density range. Marker gene expression across subtypes was displayed as dot plots; expression values were standardized per gene across groups (standard_scale = ‘var’), with dot size encoding the fraction of cells expressing each gene and dot color encoding standardized mean expression.

### Pairwise spatial co-localization enrichment analysis

Spatial proximity between cell types was assessed using a permutation-based neighborhood enrichment framework applied to each sample independently. For each sample, a spatial neighbor graph was constructed using Squidpy^60^ (sq.gr.spatial_neighbors; coord_type=’generic’). Observed pairwise interaction counts between all cell-type combinations were derived from the resulting adjacency matrix. To generate a null distribution, cell-type labels were randomly permuted across the fixed spatial graph for 5,000 iterations per sample, preserving tissue geometry while breaking cell-type spatial structure. The permutation mean served as the expected count for each cell-type pair. Interaction enrichment was quantified as log2(observed + 1) / (expected + 1), where the pseudocount of 1 was added to both terms to stabilize low-count pairs. Empirical two-sided p-values were computed as p = 2 × min(p_upper, p_lower), with a continuity-corrected formulation, p_upper = (number of permutations ≥ observed + 1) / (n_perms + 1), and were corrected for multiple testing across all cell-type pairs per sample using BH-FDR (statsmodels.stats.multitest.fdrcorrection; α = 0.05)^65^. Cell-type pairs with a permutation-expected count below 5 were excluded from significance assessment.

Per-sample and consensus results were visualized as three-dimensional bar charts using Plotly^66^. Each cell-type pair is represented as a rectangular bar positioned on a source × neighbor grid; bar height encodes the absolute value of log2(observed/expected), and bar color encodes the signed value through a diverging RdBu_r colormap (red, enriched; blue, depleted), normalized with a two-slope scale centered at zero and spanning the empirical data range. For the consensus visualization, bar heights and colors reflect the mean log2(observed/expected) values, averaged across all ten samples. Select immune-neuroblast cell-type interactions of interest are highlighted by a circle drawn on the top face of the corresponding bar.

### Top ten spatial neighbor composition analysis

To characterize the immediate cellular microenvironment of each T-cell subtype, the spatial neighbor composition was quantified for each sample. For each sample, a spatial neighbor graph was constructed using Squidpy (sq.gr.spatial_neighbors; coord_type=’generic; n_neighs=10). For each of the five T-cell subtypes (γδT, Treg, Naive/CM T, Proliferating T, Cytotoxic T), the broad cell-type labels of all graph-adjacent neighbors were collected across all T cells of that subtype. Cell-type proportions were computed as normalized frequencies across the pooled neighbor set. T-cell subtypes with fewer than ten cells in a given sample were excluded from the analysis of that sample. A consensus neighbor composition profile was derived by averaging per-sample proportions across all samples with equal weighting. Neighbor composition was visualized as stacked bar plots per T-cell subtype, with neighbor cell types ranked by mean proportion across subtypes.

### Tissue domain inference and spatial stratification of T-cell states

Compositionally distinct tissue domains were identified using Scimap^26^. For each cell, the counts of neighboring cell subtypes (from the celltypes_all annotation) within an 80 µm radius were computed using sm.tl.spatial_count (method=’radius’; radius=80), applied jointly across all ten samples. The resulting per-cell neighborhood composition profiles were clustered using k-means (sm.tl.spatial_cluster; k=20; random seed = 26), yielding 20 spatial domains. Domains were subsequently classified into three macro-categories: immune-rich (domains with total immune cell proportion ≥ 0.5, comprising macrophages, T cells, and B cells), neuroblast-rich (neuroblast proportion ≥ 0.5), and other.

To assess T-cell subtype distribution across spatial contexts, within-domain T-cell subtype proportions were computed per sample as the fraction of T cells in each domain assigned to each subtype. For each T-cell subtype, domain dependence was first evaluated using a Friedman test across the three domain categories, restricted to samples in which all three domains were represented (minimum n = 3). Pairwise *post hoc* comparisons between domain categories were performed using two-sided paired Wilcoxon signed-rank tests (scipy.stats.wilcoxon; zero_method=’wilcox’; exact method where feasible), matched by sample. BH-FDR correction was applied separately across T-cell subtypes for the Friedman omnibus tests, and across all pairwise comparisons for the Wilcoxon *post hoc* tests (statsmodels.stats.multitest.multipletests).

To compare T-cell transcriptional states across spatial contexts, pseudobulk mean gene expression was computed separately for immune-rich and neuroblast-rich domains. Genes spanning lineage (*CD4*, *CD8A*, *TRGC2*), naive/central memory (*CCR7*, *SELL*, *LEF1*, *TCF7*, *IL7R*), proliferation (*MKI67*), cytotoxic (*FOS*, *IFNG*, *GZMH*, *GZMB*, *GZMK*, *KLRD1*), and inhibitory (*PDCD1*, *LAG3*, *TOX*, *TIGIT*) programs were tested using two-sided paired Wilcoxon signed-rank tests (minimum n = 3 sample pairs), with BH-FDR correction across all genes.

### Spatial TCR clonotype analysis

Clonotype-positive cells were identified in the single-cell Xenium AnnData by summing raw probe counts across all probes sharing a clone prefix and applying a nonzero threshold. For αβ clonotype pairs, cells were required to be double-positive (carrying nonzero counts for both the α-chain and β-chain probe sets) within the same cell. Chain concordance was enforced at detection: γδ clonotype calls required concurrent nonzero expression of *TRGC2* and *CD3E*; αβ CD8 calls required *CD8A* and *CD3E*; αβ *CD4* calls required *CD4* and *CD3E*. Clonotype fraction per sample was computed as the number of clone-positive T cells divided by total T cells in that sample. Longitudinal dynamics were classified using the resection/biopsy fraction ratio (expanded > 1.10; contracted < 0.90; stable 0.90-1.10).

Differential expression between clone-positive and all other T cells within the same sample was performed using sc.tl.rank_genes_groups (Wilcoxon rank-sum test), with clone probe features excluded from the tested gene set. A minimum of 3 clone-positive cells was required per sample. To assess neoantigen-reactive T cell features, gene signature scores for four published neoantigen-reactive CD8+ TIL programs (Caushi, Oliveira, Lowery, Hanada)^46–50^ were computed using sc.tl.score_genes, restricted to genes present in the Xenium panel. Scores were compared between clonotype-positive and clonotype-negative T cells within the same sample using two-sided Mann-Whitney U tests (scipy.stats.mannwhitneyu), with BH-FDR correction applied jointly across all pairs, signatures, and timepoints (statsmodels.stats.multitest.fdrcorrection).

Domain-stratified gene expression in clone-positive cells was assessed using a stratified permutation test. For each clone-gene-domain-pair combination, the observed test statistic was the mean across samples of the within-sample expression difference (domain 2 mean - domain 1 mean). Domain labels were then permuted within each sample (10,000 permutations), and an empirical two-sided p-value was computed as (number of permuted statistics ≥ |observed| + 1) / (n_perms + 1). Only samples contributing a minimum of 10 clone-positive cells to each of the two domains being compared were included. Gene expression was visualized as dot plots with per-gene standardization across domain categories (standard_scale = ‘var’).

### Data and code availability

Raw CapTCR-seq reads and MiXCR outputs from patients enrolled in the ANBL1531 cohort are being uploaded to the database of Genotypes and Phenotypes (dbGaP). Xenium spatial transcriptomic data is available upon request. All codes used to generate figures and perform analyses and their corresponding source data are deposited on GitHub (https://github.com/pughlab/neuroblastoma_spatial_tcr).

## Data Availability

Raw CapTCR-seq reads and MiXCR outputs in the present study are being uploaded to the database of Genotypes and Phenotypes (dbGaP). Xenium spatial transcriptomic data in the present study is available upon request.

## Acknowledgements

We thank the patients and their families for their participation. We also acknowledge the contributions of the BPC staff. This work was primarily supported by the Department of Defense Congressionally Directed Medical Research Programs through awards GRANT13451092 (T.J.P.), GRANT13450969 (T.K.) and GRANT13450947 (S.D.). Biological specimens were obtained through the ANBL1531 trial (NCT03126916) supported by the NCTN Operations Center Grant U10CA180886 and the NCTN Statistics & Data Center Grant U10CA180899 from the St. Baldrick’s Foundation. Additional support was provided by the Princess Margaret Cancer Foundation, U54 CA274516, Soupy for Loopy Foundation, Sammy’s Superheroes Foundation, Cody’s Crew, and Santos Foundation. This research was enabled in part by computational resources and support provided by HPC4Health and Compute Canada. We acknowledge the Children’s Hospital of Philadelphia Center for Childhood Cancer Research Biobank, Pathology Core, Single Cell Technology Core, and Research Information Services for providing support.

T.J.P. holds the Canada Research Chair in Translational Genomics and is also supported by a Senior Investigator Award from the Ontario Institute for Cancer Research (OICR), the OICR Investigator Award (IA-031), funded by the Government of Ontario, and the Princess Margaret Cancer Foundation. KT holds the Richard and Sheila Sanford Endowed Chair at CHOP. Y.J. is additionally supported by a Canada Graduate Research Scholarship (Doctoral) from the Canadian Institutes of Health Research (FBD# 199461).

## Additional Information

The content is solely the responsibility of the authors and does not necessarily represent the official views of the National Institutes of Health.

## Figure Legends

**Extended Data Fig. 1.**
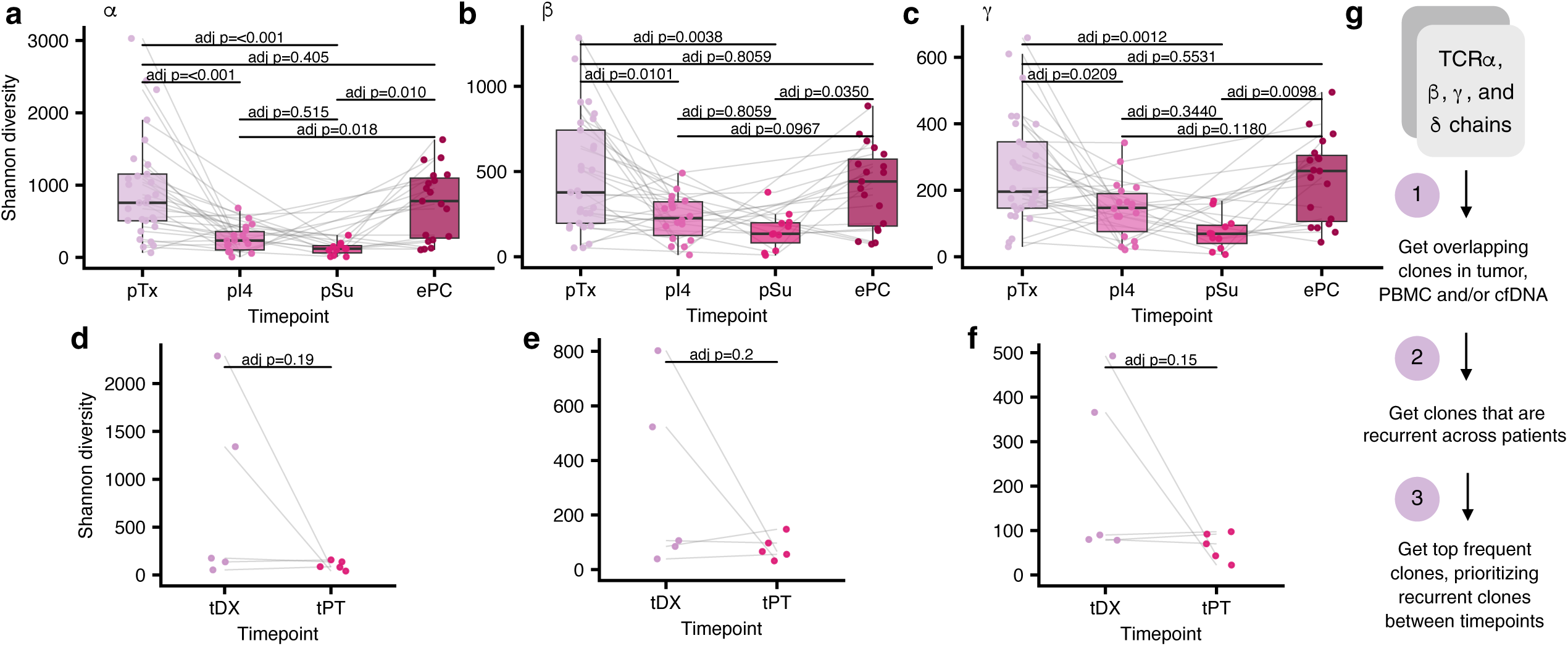
Bulk TCR repertoire dynamics and CDR3 probe selection for Xenium spatial profiling. a-c, TCR repertoire Shannon diversity for PBMC samples across four timepoints (pTx, n = 28; pI4, n = 12; pSu, n = 11; ePC, n = 18) for TCR⍺ (a), TCRβ (b) and TCRγ (c) chains. In a-c, boxes show the median and interquartile range; points represent individual samples connected by patient lines. Pairwise comparisons from a linear mixed model with patient as random effect; Holm-adjusted P values are indicated on each panel. d-f, Same metrics as a-c for paired tumor samples at DX and PT (n = 5 patients, n = 10 samples). g, Summary of the three criteria applied to select the 100 candidate CDR3 sequences for Xenium probe design: concordant clonotype abundance across tumor and PBMC compartments, high abundance within at least one specimen, and representation of candidate αβ chain-paired clonotypes and recurrent γ-chain sequences across patients.

**Extended Data Fig. 2.**
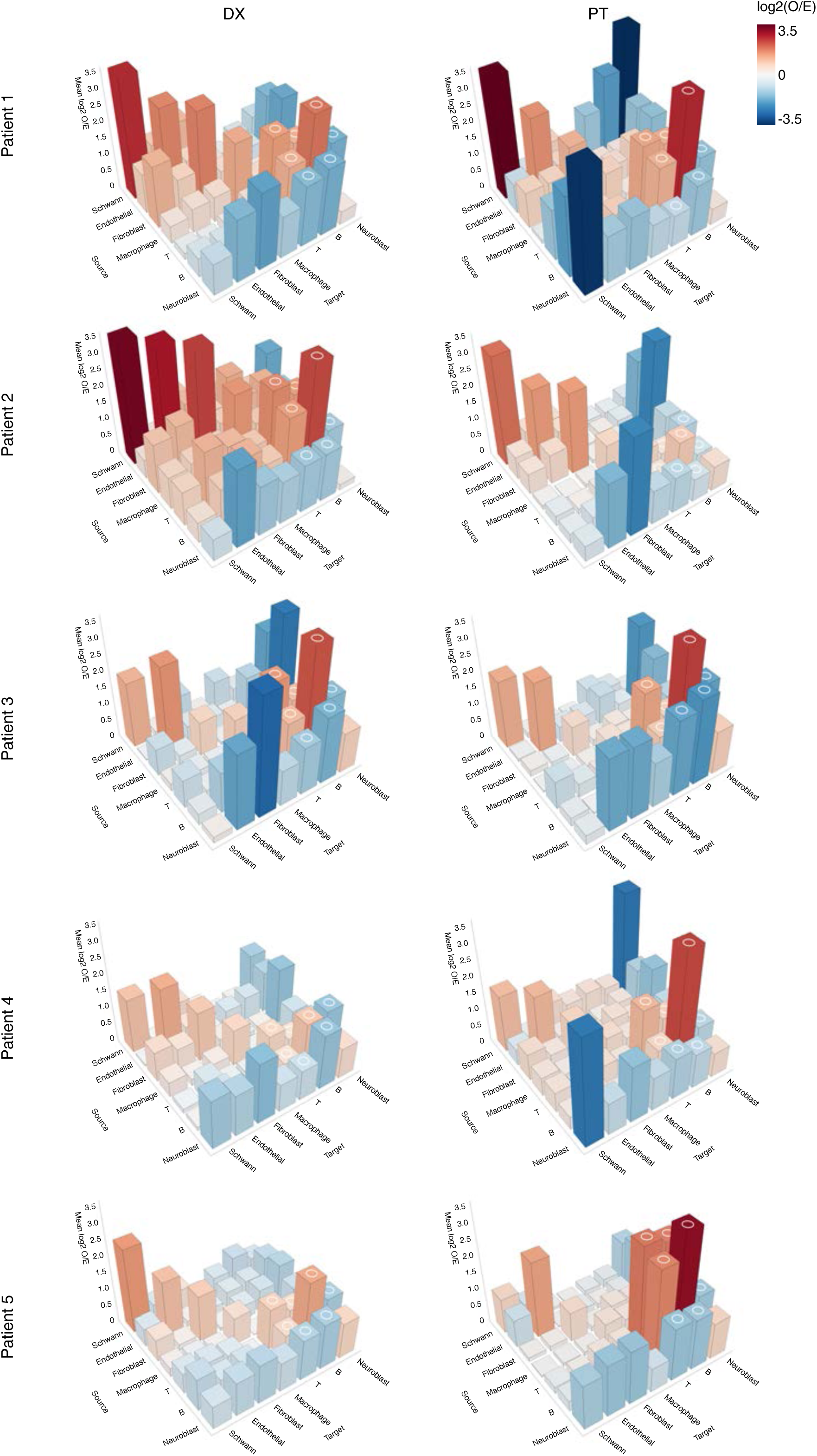
Per-sample pairwise distance enrichment 3D barplots. 3D barplots showing pairwise co-localization enrichment for each of the ten FFPE samples, computed as described for Fig. 3c. Circles denote reciprocal B-T, T-T, B-B, T-neuroblast, and B-neuroblast interactions concordantly enriched or depleted across all ten samples.

**Extended Data Fig. 3.**
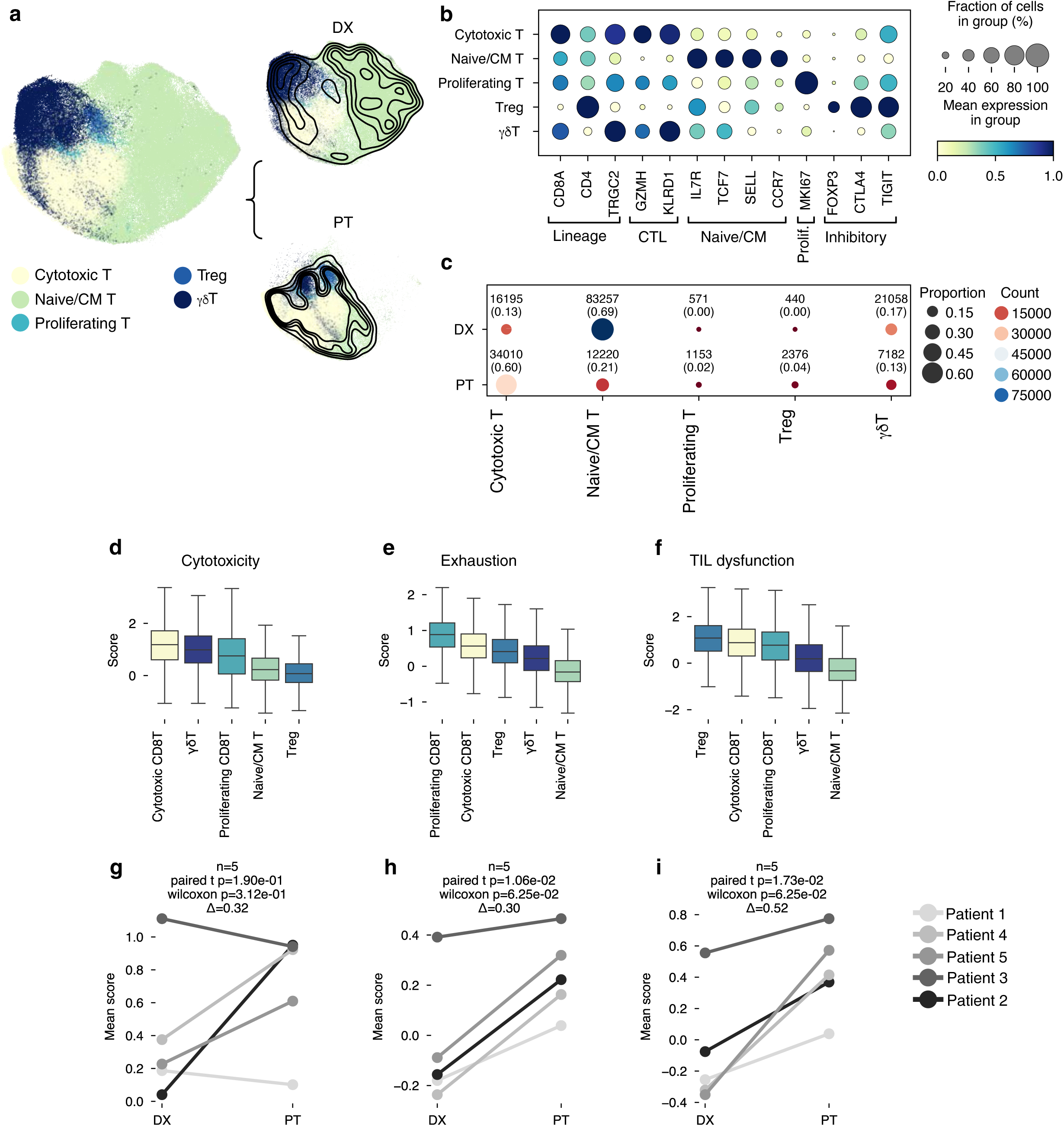
T-cell subtyping and functional gene-signature scores across subtypes and timepoints. **a**, UMAP of 178,462 T cells colored by five annotated subtypes (left), and the same embedding stratified by time point (DX and PT, right) with Gaussian kernel density contour overlays. **b**, Dot plot of lineage, cytotoxic, naive/central memory, proliferating, and inhibitory T-cell markers across subtypes. Dot size, fraction expressing; color, standardized mean expression. **c**, Dot plot showing T-subtype composition by time point. Dot size, subtype fraction for each time point; color, absolute cell count for each time point. **d**-**f**, Box plots of cytotoxicity (**d**), exhaustion (**e**), and TIL dysfunction (**f**) scores across the five T-cell subtypes (n = 178,462 cells). Scores were computed from published gene signatures. Subtypes are ordered by decreasing median. Boxes show the median and interquartile range; whiskers extend to 1.5× of the interquartile range; outliers not shown. Subtype cell counts: Naive/CM T, n = 95,477; Cytotoxic T, n = 50,205; γδT, n = 28,240; Treg, n = 2,816; Proliferating T, n = 1,724. **g**-**i**, Paired patient-level point plots of pseudobulk mean cytotoxicity (**g**), exhaustion (**h**), and TIL dysfunction (**i**) scores at DX and PT (n = 5 patient pairs). Lines connect the matched DX and PT samples for each patient. P values were computed using a two-sided paired t-test and adjusted across the three comparisons using the Benjamini-Hochberg method. Cytotoxicity: mean Δ (PT - DX) = 0.32, FDR-adjusted P = 0.19. Exhaustion: mean Δ = 0.30, FDR-adjusted P = 0.026. TIL dysfunction: mean Δ = 0.52, FDR-adjusted P = 0.026.

**Extended Data Fig. 4.**
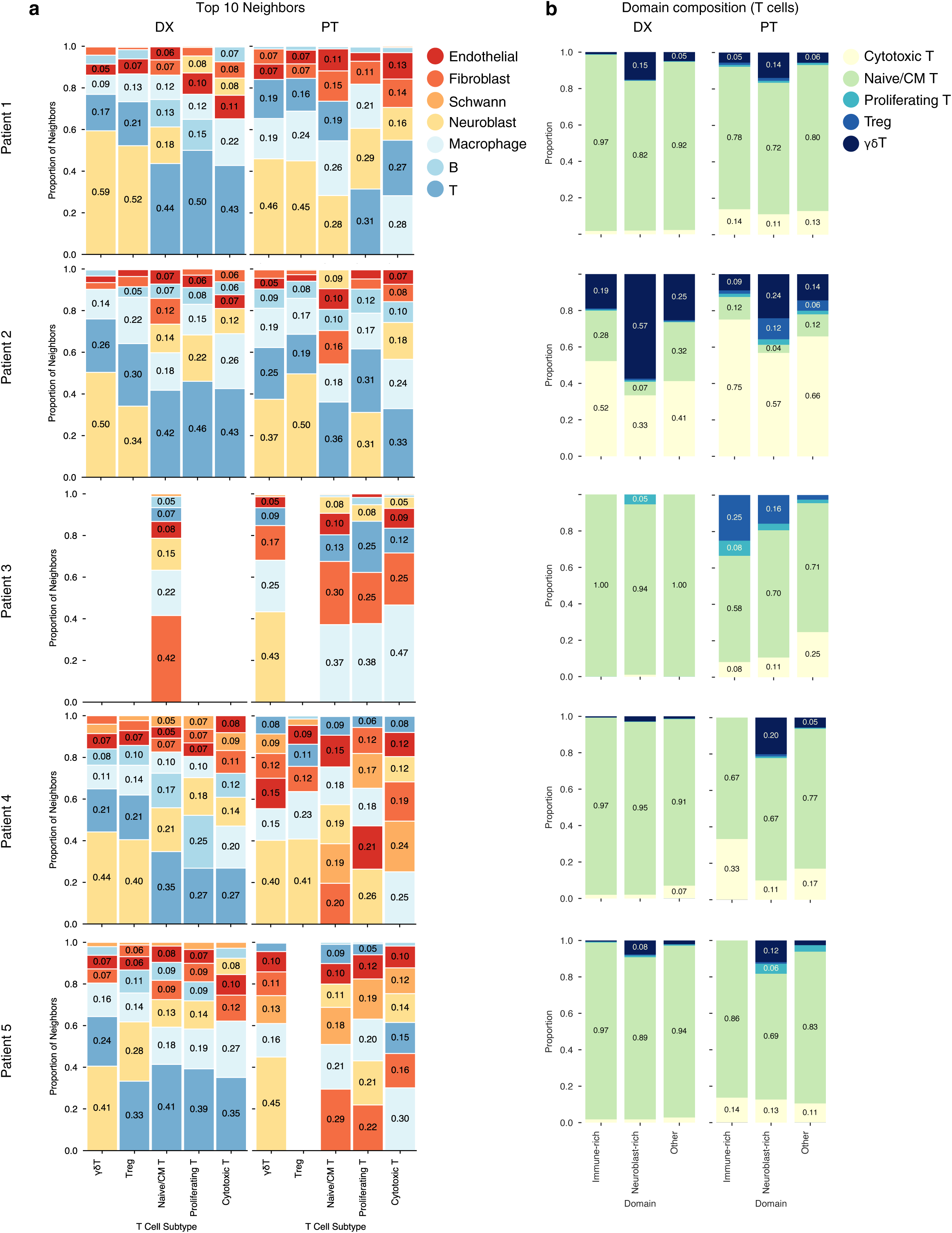
Per-sample T-cell proximity composition and domain distribution. **a**, Per-sample stacked bar plots of the major cell-type composition of the ten nearest cells to each T-cell subtype (n = 10 samples). Computed as described for Fig. 4a. Subtypes with fewer than ten cells per sample were omitted for that sample. **b**, Per-sample stacked bar plots of T-cell subtype composition across the three spatial domain categories, computed as described for Fig. 4c.

**Extended Data Fig. 5.**
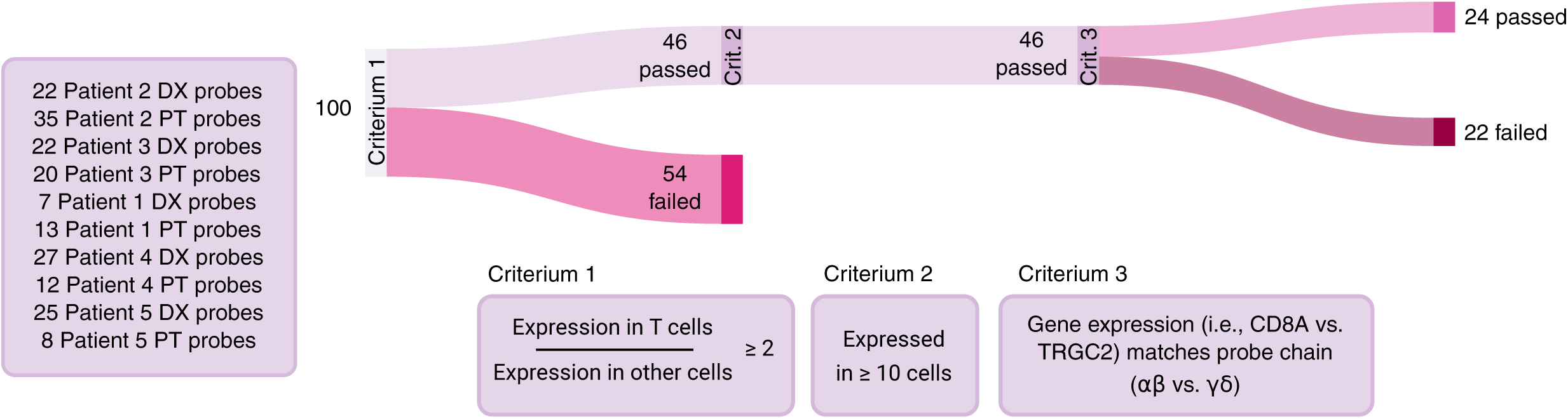
*Post hoc* CDR3 probe filtering for Xenium spatial profiling. Left, composition of the 100 candidate CDR3 probes by specimen of origin. Right top, Sankey diagram of the *post hoc* Xenium probe quality-filtering workflow: of 100 probes, 46 met T-cell enrichment and minimum detection thresholds; all 46 advanced to chain concordance assessment, of which 24 satisfied all criteria. Right bottom, the three filtering criteria applied: (i) ≥2-fold enrichment in T cells relative to non-T cells within the intended patient; (ii) detection in a minimum of 10 T cells; (iii) concordant expression with chain-specific markers (CD3E with CD4 or CD8A for TCRα/β; CD3E with TRGC2 for TCRγ/δ).

**Extended Data Fig. 6.**
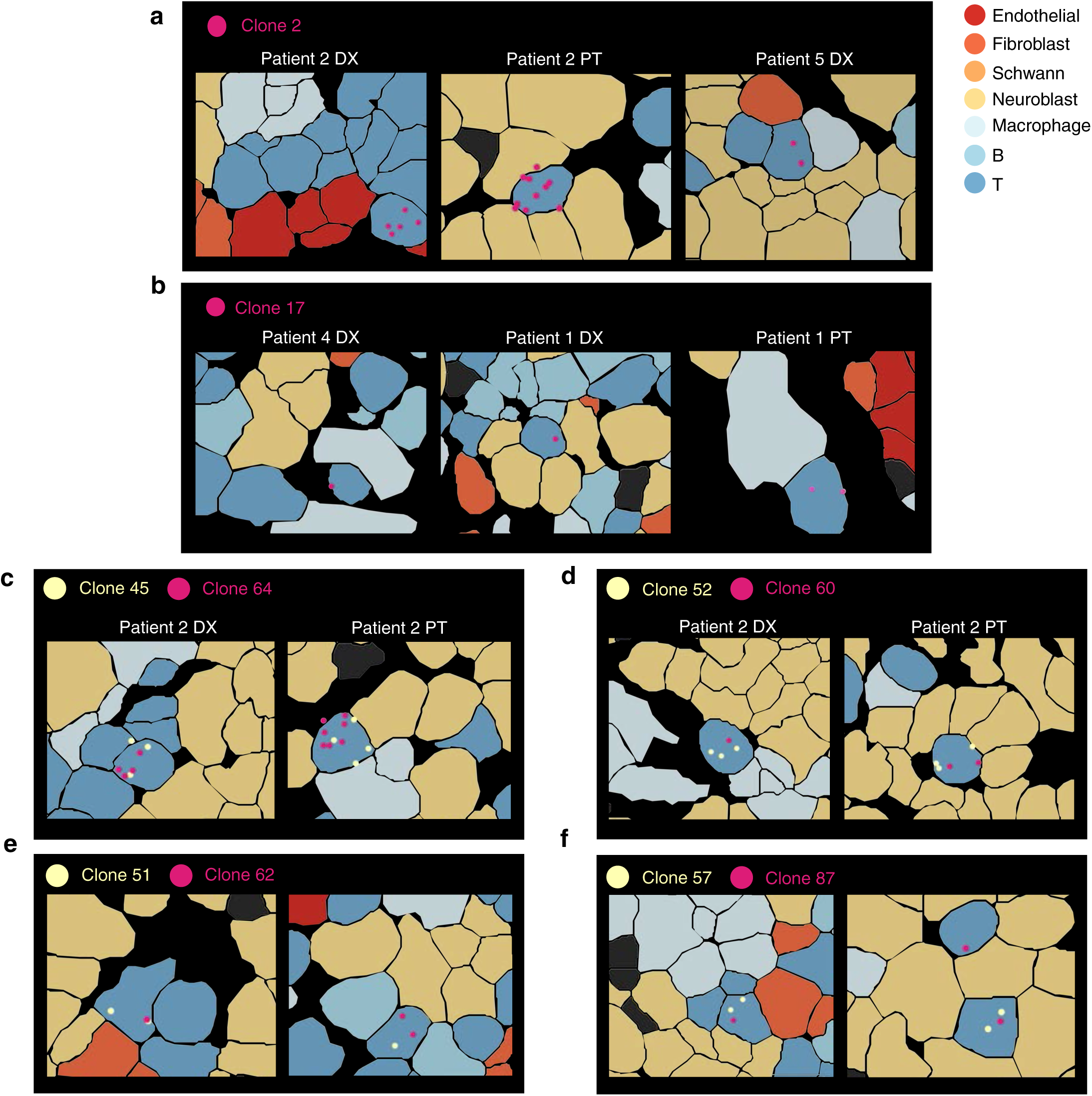
Spatial detection of validated CDR3 clonotypes and *in situ* αβ pair co-detection. **a**, Representative Xenium Explorer images of γ Clone 2 at single-cell resolution in Patient 2 DX, Patient 2 PT, and Patient 5 DX samples, showing individual CDR3 transcript detections. Each dot represents a TCR transcript. **b**, Representative Xenium Explorer images of γ Clone 17 in Patient 4 DX, Patient 1 DX, and Patient 1 PT samples, as in **a**. **c**-**f**, Representative Xenium Explorer images showing single-cell co-detection of paired CDR3 probes: αβ Pair 1 (**c**, α clone 45 + β clone 64), αβ Pair 2 (**d**, α clone 52 + β clone 60), αβ Pair 3 (**e**, α clone 51 + β clone 62) and αβ Pair 4 (**f**, α clone 57 + β clone 87), in Patient 2 DX and PT samples.

**Extended Data Fig. 7.**
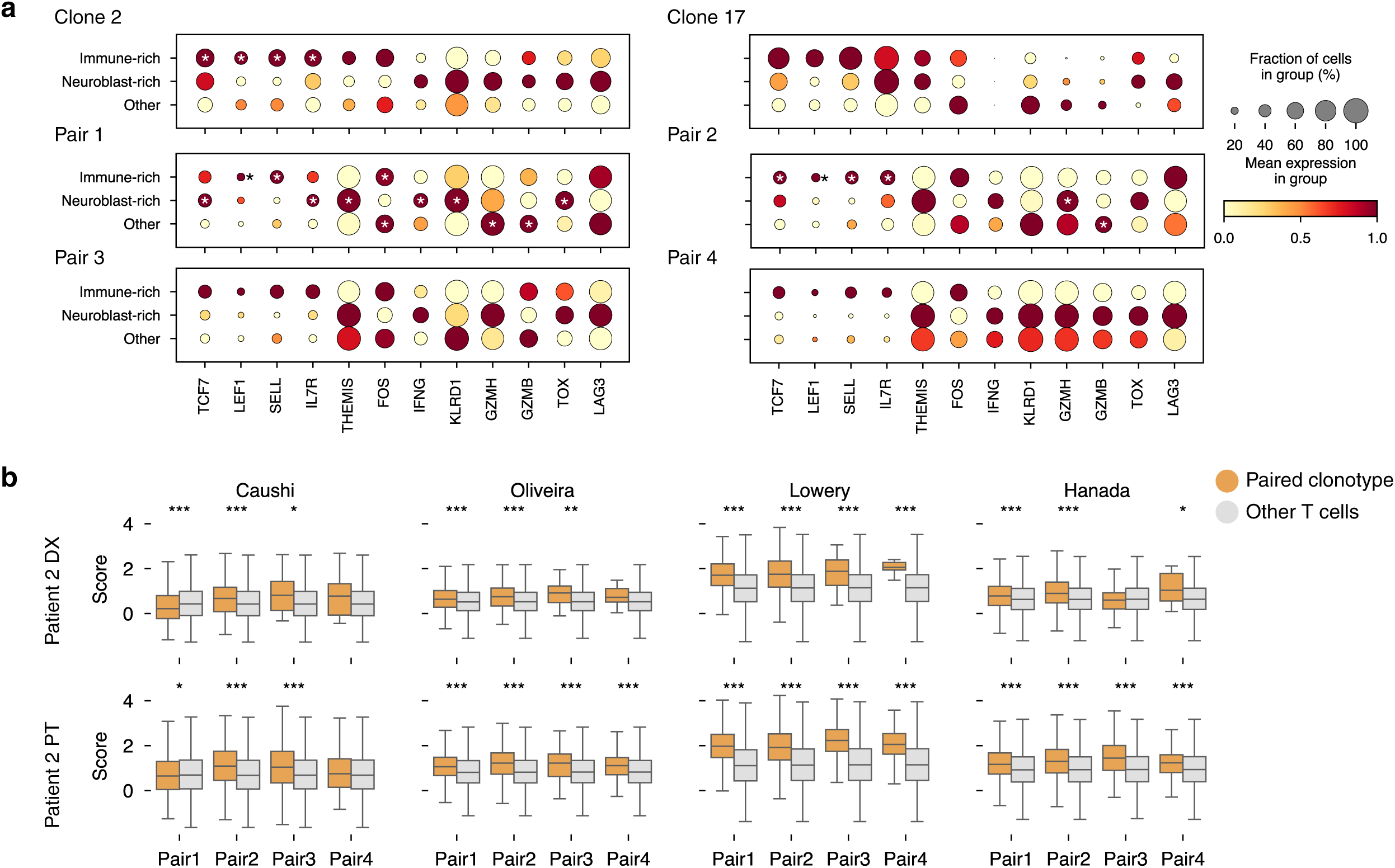
Pair-positive T cells show domain-associated transcriptional bifurcation and neoantigen-reactive TIL signature enrichment **a**, Dot plot of marker expression in γ Clone 2-, Clone 17-, Pair 1-, Pair 2-, Pair 3-, and Pair 4-positive T cells stratified by spatial domain (immune-rich, neuroblast-rich, other). Markers span naive/central memory (*TCF7*, *LEF1*, *SELL*, *IL7R*), TCR signaling (*THEMIS*), activation and cytotoxic effector (*FOS*, *IFNG*, *KLRD1*, *GZMH*, *GZMB*) and dysfunction-associated (*TOX*, *LAG3*) programs. Dot size, fraction expressing; color, standardized mean expression. *, FDR < 0.05. **b**, Box plots comparing published neoantigen-reactive TIL gene-signature scores between clonotype pair-positive and all other T cells in Patient 2, for each of the four αβ pairs at DX (top row) and PT (bottom row). Four signatures are shown in separate columns: Caushi, Oliveira, Lowery, and Hanada, each restricted to genes present on the Xenium panel. Boxes show median and interquartile range; outliers not shown. Stars indicate BH-FDR-adjusted significance from a two-sided Mann-Whitney U test: *, FDR < 0.05; **, FDR < 0.01; ***, FDR < 0.001.

## References

1. Yue, C., Zhang, Q., Sun, F. & Pan, Q. Global, regional and national burden of neuroblastoma and other peripheral nervous system tumors, 1990 to 2021 and predictions to 2035: visualizing epidemiological characteristics based on GBD 2021. Neoplasia 60, 101122 (2025).

2. Johnsen, J. I., Dyberg, C. & Wickström, M. Neuroblastoma—A neural crest derived embryonal malignancy. Front. Mol. Neurosci. 12, 9 (2019).

3. Irwin, M. S. et al. Revised neuroblastoma risk classification system: a report from the Children’s Oncology Group. J. Clin. Oncol. 39, 3229–3241 (2021).

4. Krystal, J. & Foster, J. H. Treatment of high-risk neuroblastoma. Children (Basel) 10, 1302 (2023).

5. Williams, K. M. & Gress, R. E. T cell immune reconstitution following lymphodepletion. Semin. Immunol. 19, 318–330 (2007).

6. King, E., Struck, R. & Piskareva, O. The triad in current neuroblastoma challenges: targeting antigens, enhancing effective cytotoxicity and accurate 3D in vitro modelling. Transl. Oncol. 51, 102176 (2025).

7. DuBois, S. G., Macy, M. E. & Henderson, T. O. High-risk and relapsed neuroblastoma: toward more cures and better outcomes. Am. Soc. Clin. Oncol. Educ. Book 42, 1–13 (2022).

8. Maggi, E., Landolina, N., Munari, E., Mariotti, F. R., Tumino, N., Vacca, P., Azzarone, B. & Moretta, L. T cells in the microenvironment of solid pediatric tumors: the case of neuroblastoma. Front. Immunol. 16, 1544137 (2025).

9. Schatz, D. G. & Ji, Y. Recombination centres and the orchestration of V(D)J recombination. Nat. Rev. Immunol. 11, 251–263 (2011).

10. Bao, W., Song, Z., Wan, H., Yu, X., Chen, Z., Jiang, Y., Chen, X. & Le, K. Model for predicting prognosis and immunotherapy based on CD+8 T cells infiltration in neuroblastoma. J. Cancer Res. Clin. Oncol. 149, 9839–9855 (2023).

11. Hu, Y., Hu, Q., Li, Y. et al. γδ T cells: origin and fate, subsets, diseases and immunotherapy. Sig. Transduct. Target. Ther. 8, 434 (2023).

12. Kennedy, P. T., Zannoupa, D., Son, M. H. & Dahal, L. N. Neuroblastoma: an ongoing cold front for cancer immunotherapy. J. Immunother. Cancer 11, e007798 (2023).

13. Kayano, D. & Kinuya, S. Current consensus on I-131 MIBG therapy. Nucl. Med. Mol. Imaging 52, 254–265 (2018).

14. Yu, W., Biyik-Sit, R., Uzun, Y. et al. Longitudinal single-cell multiomic atlas of high-risk neuroblastoma reveals chemotherapy-induced tumor microenvironment rewiring. Nat. Genet. 57, 1142–1154 (2025).

15. Patel, A. G. et al. A spatial cell atlas of neuroblastoma reveals developmental, epigenetic and spatial axis of tumor heterogeneity. Preprint at 10.1101/2024.01.07.574538 (2024).

16. Jansky, S., Sharma, A. K., Körber, V., Quintero, A., Toprak, U. H., Wecht, E. M., Gartlgruber, M., Greco, A., Chomsky, E., Grünewald, T. G. P., Henrich, K. O., Tanay, A., Herrmann, C., Höfer, T. & Westermann, F. Single-cell transcriptomic analyses provide insights into the developmental origins of neuroblastoma. Nat. Genet. 53, 683–693 (2021).

17. Dong, R., Yang, R., Zhan, Y., Lai, H. D., Ye, C. J., Yao, X. Y., Luo, W. Q., Cheng, X. M., Miao, J. J., Wang, J. F., Liu, B. H., Liu, X. Q., Xie, L. L., Li, Y., Zhang, M., Chen, L., Song, W. C., Qian, W., Gao, W. Q., Tang, Y. H., Shen, C. Y., Jiang, W., Chen, G., Yao, W., Dong, K. R., Xiao, X. M., Zheng, S., Li, K. & Wang, J. Single-cell characterization of malignant phenotypes and developmental trajectories of adrenal neuroblastoma. Cancer Cell 38, 716–733.e6 (2020).

18. Verhoeven, B. M., Mei, S., Olsen, T. K., Gustafsson, K., Valind, A., Lindström, A., Gisselsson, D., Fard, S. S., Hagerling, C., Kharchenko, P. V., Kogner, P., Johnsen, J. I. & Baryawno, N. The immune cell atlas of human neuroblastoma. Cell Rep. Med. 3, 100657 (2022).

19. McCord, K. A., Kan, E., Hyslop, S., Xia, A. Y., Hofferek, C. J., Lewis, J. S. Jr, Wieland, A., Hernandez, D. J., Sandulache, V. C. & Hudson, W. H. Single-cell TCR mapping reveals spatially coordinated T cell states in head and neck cancer. Sci. Immunol. 11, eaec3133 (2026).

20. Mulder, D. T., Mahé, E. R., Dowar, M., Hanna, Y., Li, T., Nguyen, L. T., Butler, M. O., Hirano, N., Delabie, J., Ohashi, P. S. & Pugh, T. J. CapTCR-seq: hybrid capture for T-cell receptor repertoire profiling. Blood Adv. 2, 3506–3514 (2018).

21. Hockett, R. D., de Villartay, J. P., Pollock, K., Poplack, D. G., Cohen, D. I. & Korsmeyer, S. J. Human T-cell antigen receptor (TCR) delta-chain locus and elements responsible for its deletion are within the TCR alpha-chain locus. Proc. Natl Acad. Sci. U.S.A. 85, 9694–9698 (1988).

22. Chen, Y.-T., Hsu, H.-C., Lee, Y.-S., Liu, H., Tan, B. C. M., Chin, C.-Y., Chang, I. Y. F. & Yang, C.-Y. Longitudinal high-throughput sequencing of the T-cell receptor repertoire reveals dynamic change and prognostic significance of peripheral blood TCR diversity in metastatic colorectal cancer during chemotherapy. Front. Immunol. 12, 743448 (2022).

23. Korsunsky, I., Millard, N., Fan, J., Slowikowski, K., Zhang, F., Wei, K., Baglaenko, Y., Brenner, M., Loh, P. & Raychaudhuri, S. Fast, sensitive and accurate integration of single-cell data with Harmony. Nat. Methods 16, 1289–1296 (2019).

24. Traag, V. A., Waltman, L. & van Eck, N. J. From Louvain to Leiden: guaranteeing well-connected communities. Sci. Rep. 9, 5233 (2019).

25. Bonine, N. et al. NBAtlas: a harmonized single-cell transcriptomic reference atlas of human neuroblastoma tumors. Cell Rep. 43, 114804 (2024).

26. Verhoeven, B. M., Mei, S., Olsen, T. K., Gustafsson, K., Valind, A., Lindström, A., Gisselsson, D., Shirazi Fard, S., Hagerling, C., Kharchenko, P. V., Kogner, P., Johnsen, J. I. & Baryawno, N. The immune cell atlas of human neuroblastoma. Cell Rep. Med. 3, 100657 (2022).

27. Wienke, J. et al. Integrative analysis of neuroblastoma by single-cell RNA sequencing identifies the NECTIN2-TIGIT axis as a target for immunotherapy. Cancer Cell 42, 283–300.e8 (2024).

28. Saura-Esteller, J., de Jong, M., King, L. A., Ensing, E., Winograd, B., de Gruijl, T. D., Parren, P. W. H. I. & van der Vliet, H. J. Gamma Delta T-Cell Based Cancer Immunotherapy: Past-Present-Future. Front. Immunol. 13, 915837 (2022).

29. Schilbach, K., Frommer, K., Meier, S., Handgretinger, R. & Eyrich, M. Immune response of human propagated gammadelta-T-cells to neuroblastoma recommend the Vdelta1+ subset for gammadelta-T-cell-based immunotherapy. J. Immunother. 31, 896–905 (2008).

30. Sarkar, T., Dhar, S. & Sa, G. Tumor-infiltrating T-regulatory cells adapt to altered metabolism to promote tumor-immune escape. Curr. Res. Immunol. 2, 132–141 (2021).

31. Nirmal, A. J. & Sorger, P. K. SCIMAP: A Python Toolkit for Integrated Spatial Analysis of Multiplexed Imaging Data. J. Open Source Softw. 9, 6604 (2024).

32. Yin, P. et al. Enhanced FOS expression improves tumor clearance and resists exhaustion in NR4A3-deficient CAR T cells under chronic antigen exposure. Sci. Adv. 11, eadw3571 (2025).

33. Castenmiller, S. M. et al. γδ T cells are the prime antitumoral T cells in pediatric neuroblastoma. Life Sci. Alliance 8, e202503249 (2025).

34. ImmunoMind Team. (2019). immunarch: An R Package for Painless Bioinformatics Analysis of T-Cell and B-Cell Immune Repertoires. Zenodo. 10.5281/zenodo.3367200

35. Shorer, O., Pinhasi, A. & Yizhak, K. Single-cell meta-analysis of T cells reveals clonal dynamics of response to checkpoint immunotherapy. Cell Genom. 5, 100842 (2025).

36. Mazet, J. M., Mahale, J. N., Tong, O., Watson, R. A., Lechuga-Vieco, A. V., Pirgova, G., Lau, V. W. C., Attar, M., Koneva, L. A., Sansom, S. N., Fairfax, B. P. & Gérard, A. IFNγ signaling in cytotoxic T cells restricts anti-tumor responses by inhibiting the maintenance and diversity of intra-tumoral stem-like T cells. Nat. Commun. 14, 321 (2023).

37. Dong, C., Lin, Z., Hu, Y. & Lu, Q. KLRD1 (CD94): A prognostic biomarker and therapeutic candidate in head and neck squamous cell carcinoma. J. Cancer 16, 982–995 (2025).

38. Weng, N.-P., Araki, Y. & Subedi, K. The molecular basis of the memory T cell response: differential gene expression and its epigenetic regulation. Nat. Rev. Immunol. 12, 306–315 (2012).

39. Khan, O. et al. TOX transcriptionally and epigenetically programs CD8+ T cell exhaustion. Nature 571, 211–218 (2019).

40. Davey, M. S. et al. The human Vδ2+ T-cell compartment comprises distinct innate-like Vγ9+ and adaptive Vγ9− subsets. Nat. Commun. 9, 1760 (2018).

41. La, C. et al. In vivo modeling of human ãä T cell ontogeny reveals terminal deoxynucleotidyl transferase as a key regulator of type 3 Vδ2 T cell development. Cell Rep. 45, 116977 (2026).

42. Gautam, N. et al. Themis controls T cell activation, effector functions, and metabolism of peripheral CD8+ T cells. Life Sci. Alliance 6, e202302156 (2023).

43. Miller, B. C. et al. Subsets of exhausted CD8+ T cells differentially mediate tumor control and respond to checkpoint blockade. Nat. Immunol. 20, 326–336 (2019).

44. Chu, T. et al. Precursors of exhausted T cells are pre-emptively formed in acute infection. Nature 640, 782–792 (2025).

45. Chowdhury, D. & Lieberman, J. Death by a thousand cuts: granzyme pathways of programmed cell death. Annu. Rev. Immunol. 26, 389–420 (2008).

46. Li, H. et al. Distinct CD8+ T cell dynamics associate with response to neoadjuvant cancer immunotherapies. Cancer Cell 43, 757–775.e8 (2025).

47. Caushi, J. X. et al. Transcriptional programs of neoantigen-specific TIL in anti-PD-1-treated lung cancers. Nature 596, 126–132 (2021).

48. Oliveira, G. et al. Phenotype, specificity and avidity of antitumour CD8+ T cells in melanoma. Nature 596, 119–125 (2021).

49. Lowery, F. J. et al. Molecular signatures of antitumor neoantigen-reactive T cells from metastatic human cancers. Science 375, eabl5447 (2022).

50. Hanada, K. et al. A phenotypic signature that identifies neoantigen-reactive T cells in fresh human lung cancers. Cancer Cell 40, 479–493.e6 (2022).

51. Jansen, C. S. et al. An intra-tumoral niche maintains and differentiates stem-like CD8 T cells. Nature 576, 465–470 (2019).

52. Zhang, W., Brown, E.L., Usmani, A. et al. Non-invasive profiling of the tumour microenvironment with spatial ecotypes. Nature (2026).

53. Bonneville, M. & Fournié, J.-J. Sensing cell stress and transformation through Vγ9Vδ2 T cell-mediated recognition of the isoprenoid pathway metabolites. Microbes Infect. 7, 503–509 (2005).

54. Aminzadeh, S., Vidali, S., Sperl, W., Kofler, B. & Feichtinger, R. G. Energy metabolism in neuroblastoma and Wilms tumor. Transl. Pediatr. 4, 20–32 (2015).

55. Bolotin, D. A., Poslavsky, S., Mitrophanov, I., Shugay, M., Mamedov, I. Z., Putintseva, E. V. & Chudakov, D. M. MiXCR: software for comprehensive adaptive immunity profiling. Nat. Methods 12, 380–381 (2015).

56. Bolotin, D. A. et al. Antigen receptor repertoire profiling from RNA-seq data. Nat. Biotechnol. 35, 908–911 (2017).

57. R Core Team. R: A Language and Environment for Statistical Computing. R Foundation for Statistical Computing, Vienna, Austria (2025).

58. Janesick, A. et al. High resolution mapping of the tumor microenvironment using integrated single-cell, spatial and in situ analysis. Nat. Commun. 14, 8353 (2023).

59. Wolf, F. A., Angerer, P. & Theis, F. J. SCANPY: large-scale single-cell gene expression data analysis. Genome Biol. 19, 15 (2018).

60. Palla, G. et al. Squidpy: a scalable framework for spatial omics analysis. Nat. Methods 19, 171–178 (2022).

61. Marconato, L. et al. SpatialData: an open and universal data framework for spatial omics. Nat. Methods 22, 58–62 (2025).

62. Jolliffe, I. T. & Cadima, J. Principal component analysis: a review and recent developments. Philos. Trans. R. Soc. A 374, 20150202 (2016).

63. McInnes, L., Healy, J. & Melville, J. UMAP: Uniform Manifold Approximation and Projection for Dimension Reduction. Preprint at https://arXiv.org/abs/1802.03426 (2018).

64. Virtanen, P. et al. SciPy 1.0: fundamental algorithms for scientific computing in Python. Nat. Methods 17, 261–272 (2020).

65. Seabold, S. & Perktold, J. statsmodels: econometric and statistical modeling with Python. Proc. 9th Python Sci. Conf. 57–61 (2010).

66. Kruchten, N., Seier, A. & Parmer, C. An interactive, open-source, and browser-based graphing library for Python. Zenodo 10.5281/zenodo.14503524 (2026).

